# Bayesian machine learning enables discovery of risk factors for hepatosplenic multimorbidity related to schistosomiasis

**DOI:** 10.1101/2025.09.19.25336151

**Authors:** Yin-Cong Zhi, Victor Anguajibi, John B. Oryema, Betty Nabatte, Christopher K. Opio, Narcis B. Kabatereine, Goylette F. Chami

**Author notes:** **Correspondence** Dr Goylette F. Chami, Big Data Institute, University of Oxford, Oxford, United Kingdom OX3 7LF.

## Abstract

One in 25 deaths worldwide is related to liver disease, and often with multiple hepatosplenic conditions. Yet, little is understood of the risk factors for hepatosplenic multimorbidity, especially in the context of chronic infections. We present a novel Bayesian multitask learning framework to jointly model 45 hepatosplenic conditions assessed using point-of-care B-mode ultrasound for 3155 individuals aged 5-91 years within the SchistoTrack cohort across rural Uganda where chronic intestinal schistosomiasis is endemic. We identified distinct and shared biomedical, socioeconomic, and spatial risk factors for individual conditions and hepatosplenic multimorbidity, and introduced methods for measuring condition dependencies as risk factors. Notably, for gastro-oesophageal varices, we discovered key risk factors of older age, lower hemoglobin concentration, and severe schistosomal liver fibrosis. Our findings provide a compendium of risk factors to inform surveillance, triage, and follow-up, while our model enables improved prediction of hepatosplenic multimorbidity, and if validated on other systems, general multimorbidity.

## Introduction

Multimorbidity is the co-occurrence of two or more chronic conditions and is of growing global concern in existing health systems due to the lack of clinical guidelines, strains on resources and staff, and potential mismanagement/misdiagnosis of patients [1–3]. Individuals with multimorbidity are among the most socially and economically disadvantaged with greater years lived with disability and higher mortality compared to those without multimorbidity [2]. In low and middle-income countries (LMICs), over one third of individuals are affected by complex multimorbidity due to both infectious and non-communicable causes [4, 5]. Yet, there is little understanding of multimorbidity due to chronic infections in LMICs with most multimorbidity studies focusing on aging or cardiovascular diseases in high-income countries [6–8].

Schistosomiasis, caused by the intestinal species *Schistosoma mansoni*, is a major cause of hepatosplenic multimorbidity in sub-Saharan Africa. Chronic schistosome infections result in a wide range of interacting or co-occurring conditions ranging from less severe forms of liver fibrosis and organomegaly to life-threatening complications of gastro-oesophageal varices and cirrhosis [9]. More than one in every two individuals aged five years and older living in areas endemic with *S. mansoni* have been estimated to have hepatosplenic multimorbidity [9]. Yet, there is a limited understanding of the risk factors of hepatosplenic multimorbidity or, more generally, diverse hepatosplenic outcomes beyond periportal fibrosis [10]. Currently, only simple composite outcomes are studied where diverse conditions are collapsed into one indicator (periportal fibrosis [10]) or less severe conditions (diarrhoea, etc. [11]) are assessed as independent outcomes. From these singular outcome studies, current schistosome infection has been shown to be irrelevant for periportal fibrosis prediction [10], despite still being used as proxy indicator for morbidity by the World Health Organization (WHO) [12–14]. In contrast, co-infection with human immunodeficiency virus (HIV) and hepatitis B (HBV) infections have been shown to confer over two times higher odds of periportal fibrosis coupled with the past history of schistosome exposure [10]. The complex interplay of pathogenesis provides support for a renewed focus on multimorbidity and interacting conditions. It is unknown whether co-infections result in biologically-related conditions that interact to exacerbate schistosomiasis-specific conditions or cause independent co-occurring conditions due to shared socioeconomic and spatial risk factors. Life-threatening conditions related to schistosomiasis, such as gastro-oesophageal varices that are indicative of portal hypertension, remain still to be studied outside of health facilities and beyond basic demographic risk factors [15] and little is known about how multimorbidity influences schistosomiasis-related condition progression.

For schistosomiasis, the common approach of focusing narrowly on one condition at a time fails to account for all conditions currently relevant to a patient. Consequently, there is a limited understanding of risk factors shared across conditions and, critically, condition inter-dependencies, to be able to identify early indicators of severe conditions and disease progression. This oversimplified approach to multimorbidity modelling is not unique to schistosomiasis and applies more generally to recent attempts to study multimorbidity. Many studies simply identify multimorbidity as a binary indicator of two or more conditions from an arbitrary list of predefined conditions regardless of the actual condition type or interaction [2, 5, 16, 17].

Given multimorbidity arises from shared risk factors including condition inter-dependencies, we developed a Bayesian multitask learning architecture [18, 19], allowing each condition to learn from the risk factors of other conditions [20]. Graphs (or networks) were utilized to encode inter-dependencies between conditions, and parameterized to homogenize the predictions based on node connectivity. We jointly predicted 45 hepatosplenic conditions assessed through point-of-care ultrasound for 3155 individuals aged 5-91 years within the community-based cohort, SchistoTrack, in rural Uganda. The aim of this study was to employ Bayesian machine learning and multitask modelling on the 45 conditions, for discovering shared risk factors from a wide range of biomedical, sociodemographic, and spatial variables for hepatosplenic multimorbidity, in particular life-threatening severe hepatosplenic conditions, and to measure the influence of condition dependencies.

## Results

### Risk factors for hepatosplenic conditions

In our study population, 82% (2578/3155) of individuals had at least one condition, and 54% (1708/3155) with two or more conditions. Mild liver fibrosis (Niamey protocol patterns C), liver shrunkenness, and spleen enlargement were the most commonly observed (Table 1). We first identified significant risk factors for each of the individual 45 conditions as listed in Table 2.

**Table 1:** List of outcome conditions. Condition groupings are indicated by the first column. Participants can have multiple conditions from the same group except for organ measurements (left and right liver lobe, mean portal vein, and spleen length) which are mutually exclusive. In each group, the healthy status is italicized and used as reference. For liver patterns, Niamey protocol grades are indicated in brackets. Summary is over 3155 participants.

| Outcome Group | Outcome | # Participants (%) | Severity |
| --- | --- | --- | --- |
| Liver patterns | *Normal* | 2312 (73.28) | none |
|  | Unclear (B) | 15 (0.48) | mild |
|  | Feather streaks (B0) | 238 (7.54) | mild |
|  | Flying saucers, starry sky (B1) | 194 (6.15) | mild |
|  | Spider thickening (B2) | 53 (1.68) | mild |
|  | Prominent peripheral rings (C1) | 461 (14.61) | mild |
|  | Prominent pipe stems (C2) | 455 (14.41) | moderate |
|  | Ruff, portal bifurcation (D) | 185 (5.86) | severe |
|  | Patches (occluded, bright white vessels) (E) | 45 (1.43) | severe |
|  | Bird's claw (F) | 8 (0.25) | severe |
| Other abnormalities | *None* | 2922 (92.61) | none |
|  | Cirrhosis-like liver | 12 (0.38) | severe |
|  | Fatty-like liver | 64 (2.03) | mild |
|  | Chronic hepatitis or early cirrhosis | 65 (2.06) | moderate |
|  | Polycystic kidney disease | 2 (0.06) | none |
|  | Liver cysts | 4 (0.13) | none |
|  | Situs inversus | 1 (0.03) | none |
|  | Other | 47 (1.49) | unclear |
| Liver surface | *None* | 3104 (98.38) | none |
|  | Slight/serrated | 17 (0.54) | moderate |
|  | Gross/undulating | 34 (1.08) | severe |
| Caudal liver edge | *Sharp* | 2881 (91.32) | none |
|  | Rounded | 274 (8.68) | mild |
| Left liver lobe | *Normal* | 2189 (69.38) | none |
|  | Moderately enlarged | 422 (13.38) | mild |
|  | Moderately shrunken | 357 (11.32) | moderate |
|  | Severely enlarged | 84 (2.66) | mild |
|  | Severely shrunken | 103 (3.26) | moderate |
| Right liver lobe | *Normal* | 2179 (69.06) | none |
|  | Moderately enlarged | 404 (12.81) | mild |
|  | Moderately shrunken | 409 (12.96) | moderate |
|  | Severely enlarged | 51 (1.62) | mild |
|  | Severely shrunken | 112 (3.55) | severe |
| Mean portal vein | *Normal* | 2138 (67.77) | none |
|  | Moderately enlarged | 421 (13.34) | moderate |
|  | Moderately restricted | 382 (12.11) | unclear |
|  | Severely enlarged | 166 (5.26) | mild |
|  | Severely restricted | 48 (1.52) | unclear |
| Portosystemic collaterals | *Not detected* | 3121 (98.92) | none |
|  | Splenic varices | 12 (0.38) | severe |
|  | Gastro-oesophageal varices | 11 (0.35) | severe |
|  | Pancreaticoduodenal varices | 5 (0.16) | severe |
| | Entirely recanalized paraumbilical vein $\geq 3\text{mm}$ | 1 (0.03) | severe |
|  | Splenorenal shunt | 13 (0.41) | severe |
|  | Other | 1 (0.03) | severe |
| Ascites | *Not detected* | 3143 (99.62) | none |
|  | Yes | 12 (0.38) | moderate |
| Gall bladder visible | Not visible | 30 (0.95) | severe |
|  | Yes, but blocked by stone or collapsed | 115 (3.65) | none |
|  | *Yes, clearly visible* | 3010 (95.40) | none |
| Gall bladder wall | *Normal* | 2908 (92.17) | none |
|  | Thick | 247 (7.83) | severe |
| Spleen length | *Normal* | 2164 (68.59) | none |
|  | Moderately enlarged | 384 (12.17) | moderate |
|  | Moderately shrunken | 390 (12.36) | none |
|  | Severely enlarged | 180 (5.71) | severe |
|  | Severely shrunken | 37 (1.17) | none |

**Table 2:**
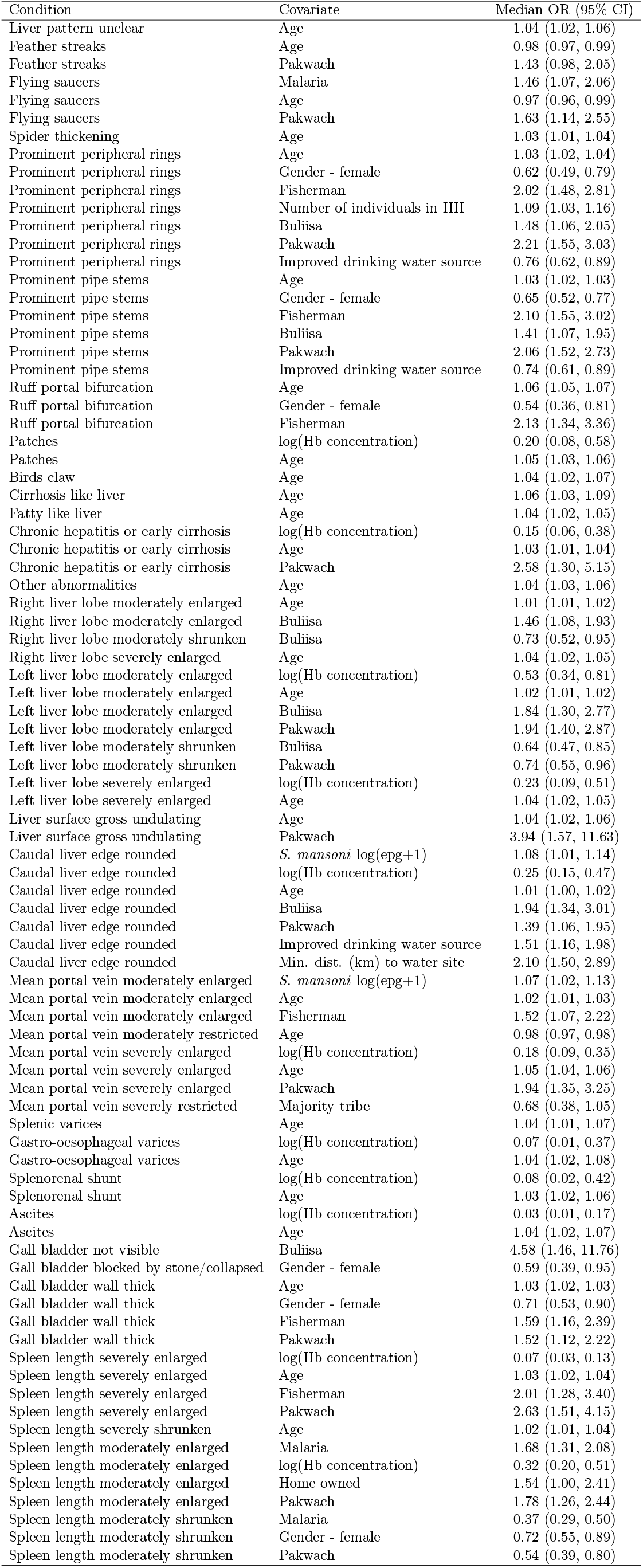
List of significant relationships found between all conditions and covariates. Outcome conditions and their significant covariates are listed in the first two columns, significance is calculated to 5% based on corrected *q*-values.

| Condition | Covariate | Median OR (95% CI) |
| --- | --- | --- |
| Liver pattern unclear | Age | 1.04 (1.02, 1.06) |
| Feather streaks | Age | 0.98 (0.97, 0.99) |
| Feather streaks | Pakwach | 1.43 (0.98, 2.05) |
| Flying saucers | Malaria | 1.46 (1.07, 2.06) |
| Flying saucers | Age | 0.97 (0.96, 0.99) |
| Flying saucers | Pakwach | 1.63 (1.14, 2.55) |
| Spider thickening | Age | 1.03 (1.01, 1.04) |
| Prominent peripheral rings | Age | 1.03 (1.02, 1.04) |
| Prominent peripheral rings | Gender - female | 0.62 (0.49, 0.79) |
| Prominent peripheral rings | Fisherman | 2.02 (1.48, 2.81) |
| Prominent peripheral rings | Number of individuals in HH | 1.09 (1.03, 1.16) |
| Prominent peripheral rings | Buliisa | 1.48 (1.06, 2.05) |
| Prominent peripheral rings | Pakwach | 2.21 (1.55, 3.03) |
| Prominent peripheral rings | Improved drinking water source | 0.76 (0.62, 0.89) |
| Prominent pipe stems | Age | 1.03 (1.02, 1.03) |
| Prominent pipe stems | Gender - female | 0.65 (0.52, 0.77) |
| Prominent pipe stems | Fisherman | 2.10 (1.55, 3.02) |
| Prominent pipe stems | Buliisa | 1.41 (1.07, 1.95) |
| Prominent pipe stems | Pakwach | 2.06 (1.52, 2.73) |
| Prominent pipe stems | Improved drinking water source | 0.74 (0.61, 0.89) |
| Ruff portal bifurcation | Age | 1.06 (1.05, 1.07) |
| Ruff portal bifurcation | Gender - female | 0.54 (0.36, 0.81) |
| Ruff portal bifurcation | Fisherman | 2.13 (1.34, 3.36) |
| Patches | log(Hb concentration) | 0.20 (0.08, 0.58) |
| Patches | Age | 1.05 (1.03, 1.06) |
| Birds claw | Age | 1.04 (1.02, 1.07) |
| Cirrhosis like liver | Age | 1.06 (1.03, 1.09) |
| Fatty like liver | Age | 1.04 (1.02, 1.05) |
| Chronic hepatitis or early cirrhosis | log(Hb concentration) | 0.15 (0.06, 0.38) |
| Chronic hepatitis or early cirrhosis | Age | 1.03 (1.01, 1.04) |
| Chronic hepatitis or early cirrhosis | Pakwach | 2.58 (1.30, 5.15) |
| Other abnormalities | Age | 1.04 (1.03, 1.06) |
| Right liver lobe moderately enlarged | Age | 1.01 (1.01, 1.02) |
| Right liver lobe moderately enlarged | Buliisa | 1.46 (1.08, 1.93) |
| Right liver lobe moderately shrunken | Buliisa | 0.73 (0.52, 0.95) |
| Right liver lobe severely enlarged | Age | 1.04 (1.02, 1.05) |
| Left liver lobe moderately enlarged | log(Hb concentration) | 0.53 (0.34, 0.81) |
| Left liver lobe moderately enlarged | Age | 1.02 (1.01, 1.02) |
| Left liver lobe moderately enlarged | Buliisa | 1.84 (1.30, 2.77) |
| Left liver lobe moderately enlarged | Pakwach | 1.94 (1.40, 2.87) |
| Left liver lobe moderately shrunken | Buliisa | 0.64 (0.47, 0.85) |
| Left liver lobe moderately shrunken | Pakwach | 0.74 (0.55, 0.96) |
| Left liver lobe severely enlarged | log(Hb concentration) | 0.23 (0.09, 0.51) |
| Left liver lobe severely enlarged | Age | 1.04 (1.02, 1.05) |
| Liver surface gross undulating | Age | 1.04 (1.02, 1.06) |
| Liver surface gross undulating | Pakwach | 3.94 (1.57, 11.63) |
| Caudal liver edge rounded | *S. mansoni* log(epg+1) | 1.08 (1.01, 1.14) |
| Caudal liver edge rounded | log(Hb concentration) | 0.25 (0.15, 0.47) |
| Caudal liver edge rounded | Age | 1.01 (1.00, 1.02) |
| Caudal liver edge rounded | Buliisa | 1.94 (1.34, 3.01) |
| Caudal liver edge rounded | Pakwach | 1.39 (1.06, 1.95) |
| Caudal liver edge rounded | Improved drinking water source | 1.51 (1.16, 1.98) |
| Caudal liver edge rounded | Min. dist. (km) to water site | 2.10 (1.50, 2.89) |
| Mean portal vein moderately enlarged | *S. mansoni* log(epg+1) | 1.07 (1.02, 1.13) |
| Mean portal vein moderately enlarged | Age | 1.02 (1.01, 1.03) |
| Mean portal vein moderately enlarged | Fisherman | 1.52 (1.07, 2.22) |
| Mean portal vein moderately restricted | Age | 0.98 (0.97, 0.98) |
| Mean portal vein severely enlarged | log(Hb concentration) | 0.18 (0.09, 0.35) |
| Mean portal vein severely enlarged | Age | 1.05 (1.04, 1.06) |
| Mean portal vein severely enlarged | Pakwach | 1.94 (1.35, 3.25) |
| Mean portal vein severely restricted | Majority tribe | 0.68 (0.38, 1.05) |
| Splenic varices | Age | 1.04 (1.01, 1.07) |
| Gastro-oesophageal varices | log(Hb concentration) | 0.07 (0.01, 0.37) |
| Gastro-oesophageal varices | Age | 1.04 (1.02, 1.08) |
| Splenorenal shunt | log(Hb concentration) | 0.08 (0.02, 0.42) |
| Splenorenal shunt | Age | 1.03 (1.02, 1.06) |
| Ascites | log(Hb concentration) | 0.03 (0.01, 0.17) |
| Ascites | Age | 1.04 (1.02, 1.07) |
| Gall bladder not visible | Buliisa | 4.58 (1.46, 11.76) |
| Gall bladder blocked by stone/collapsed | Gender - female | 0.59 (0.39, 0.95) |
| Gall bladder wall thick | Age | 1.03 (1.02, 1.03) |
| Gall bladder wall thick | Gender - female | 0.71 (0.53, 0.90) |
| Gall bladder wall thick | Fisherman | 1.59 (1.16, 2.39) |
| Gall bladder wall thick | Pakwach | 1.52 (1.12, 2.22) |
| Spleen length severely enlarged | log(Hb concentration) | 0.07 (0.03, 0.13) |
| Spleen length severely enlarged | Age | 1.03 (1.02, 1.04) |
| Spleen length severely enlarged | Fisherman | 2.01 (1.28, 3.40) |
| Spleen length severely enlarged | Pakwach | 2.63 (1.51, 4.15) |
| Spleen length severely shrunken | Age | 1.02 (1.01, 1.04) |
| Spleen length moderately enlarged | Malaria | 1.68 (1.31, 2.08) |
| Spleen length moderately enlarged | log(Hb concentration) | 0.32 (0.20, 0.51) |
| Spleen length moderately enlarged | Home owned | 1.54 (1.00, 2.41) |
| Spleen length moderately enlarged | Pakwach | 1.78 (1.26, 2.44) |
| Spleen length moderately shrunken | Malaria | 0.37 (0.29, 0.50) |
| Spleen length moderately shrunken | Gender - female | 0.72 (0.55, 0.89) |
| Spleen length moderately shrunken | Pakwach | 0.54 (0.39, 0.80) |

Risk factors significant for at least one condition included three clinical measurements, seven sociodemographic variables, and three spatial factors. Hemoglobin (Hb) concentration was the most informative measurement and was negatively related to 24.44% (11/45) of conditions (natural log-transformed average median odds ratios (OR) 0.20, range 0.03 - 0.54); among associated conditions, four were severe, four moderate, and three mild with respect to portal hypertension risk. Notably, among these conditions, variation in WHO categories of anaemia status were observed for chronic hepatitis or early cirrhosis, gastro-oesophageal varices, splenorenal shunts, ascites, and severely enlarged spleens (Supplementary Table S6). Malaria infection and *S. mansoni* eggs per gram were associated with few non-severe conditions (three and two, respectively). Age was relevant for most (64.44%, 29/45) conditions irrespective of severity (10 severe, four moderate, 12 mild) with average median OR of 1.03 (range 0.97 - 1.06), and almost all significant conditions were positively related to older age (89.66%, 26/29). Both gender (female) and being a fisherman were related to six conditions (13.33%). All associations with being female were negative (average median OR 0.64, range 0.53 - 0.72) including two severe conditions, and all associations with being a fisherman were positive (average median OR 1.91, range 1.51 - 2.13) including three severe conditions. Access to an improved drinking water source had two negative (median OR range 0.74 - 0.75) and one positive (median OR 1.57) association with only non-severe conditions. The number of individuals in the household (positive), belonging to the majority tribe (negative), and belonging to a home that was owned (i.e. not rented) (positive) were related to one non-severe condition each (OR range 0.66 - 1.53). Spatial risk factors included living in the Western region of Uganda and distance to open freshwater sites. Pakwach district was associated to 31.11% (14/45, 12/14 positive) of conditions, and Buliisa district to 17.78% (8/45, 6/8 positive) when compared to Mayuge district, of which only three and one condition, respectively, were severe. The minimum distance (km) to an open freshwater site was positively associated with only one mild condition. The OR for every condition is presented in Supplementary Fig. S3, presented by the mean to show the directional effect.

Gastro-oesophageal varices were the primary focus of our analysis because they indicate severe portal hypertension. All covariate significance values for the condition are shown in Supplementary Table S3; the corresponding posterior densities are visualized in Supplementary Fig. S2. Age and Hb concentration were the only significant variables. Each additional year of age was associated with a 4% increase in the odds of gastro-oesophageal varices (median OR 1.04, 95% credible interval (CI) 1.02 - 1.08). A 10% increase in the log-transformed Hb concentration corresponded to a 22% decrease in the likelihood of gastro-oesophageal varices (OR 0.07, 95% CI 0.01 - 0.37). From the covariate densities, a number of clear moderate to large non-zero effects of other risk factors were observed, but were borderline insignificant from uncorrected *p*-values. HIV (OR 2.52, CI 0.97 - 10.77), HBV (OR 1.40, CI 0.81 - 3.72), female (OR 0.67, CI 0.26 - 1.04), and fisherman (OR 1.79, CI 0.91 - 4.44) had credible intervals narrowly including 1. The non-zero effect was visible through the heavy tails of skewed posterior densities, while insignificance was due to the modal OR remaining close to one.

Group risk factors were computed to profile periportal fibrosis associated to schistosomiasis through combined likelihood ratios of the five liver fibrosis patterns (Niamey protocol patterns C - F as defined by the features adjacent to portal vasculature) (Supplementary Table S3). Direction of association of each covariate was determined by the mean from combining the five posterior ORs densities. Periportal fibrosis shared many risk factors with the five patterns individually, with the addition of HIV despite being insignificant alone for singular liver fibrosis patterns. HIV exhibited a positive mean effect, with infection leading to a 1.77 higher odds of developing periportal fibrosis (Niamey protocol patterns C-F). Other significant associations were found with older age (mean OR 1.04), being female (mean OR 0.62), being a fisherman (mean OR 2.00), the number of individuals in the household (mean OR 1.09), access to improved drinking water source (mean OR 0.95), and the Western districts (Buliisa mean OR 1.31, Pakwach mean OR 1.99).

Significance against overall hepatosplenic multimorbidity also was computed across the 45 conditions (Supplementary Table S3). Eight risk factors previously observed to be significant for individual conditions remained, these included malaria, Hb concentration, age, being female, fisherman, distance to open freshwater sites, and the Western districts. Findings for individual conditions, periportal fibrosis, and overall hepatosplenic multimorbidity remained robust when stratified by children (aged 5-17 years) and adults (aged 18+ years), notably revealing most risk factors associations were found in adults (Supplementary Tables S1-S2, & S4-S5).

### Graph convolution and condition inter-dependencies

Condition dependencies for gastro-oesophageal varices using the graph is shown in Fig. 1 where node weightings (condition dependency measure) for the condition controlled by parameter *α* chosen as the median value from its posterior MCMC samples, (Supplementary Fig. S5). A set of probabilities between all conditions can be found in Supplementary Fig. S6. The self-influence for gastro-oesophageal varices indicated a probability of 0.397, measuring how much was predicted from its own model and risk factors/covariates. From the other condition models, the highest probability contributions to gastro-oesophageal varices came from related biological complications of splenic varices and splenorenal shunting, conferring additional probabilities of 0.102 and 0.081, respectively. The next most relevant conditions encompassed potential fibrotic precursors to portal hypertension and included moderate and severe forms of schistosomal periportal fibrosis. The ruff liver pattern (Niamey protocol pattern D) where there was extensive fibrosis at the point of portal bifurcation increased the probability by 0.045. Liver patches (Niamey protocol pattern E), which were indicative of occluded vessels and fibrosis blocking the main portal vein, conferred an increased probability of 0.059. Other condition dependencies included a severely enlarged spleen (0.049 probability), and severely enlarged (mean) portal vein diameter (0.046 probability). All other conditions had less than 0.030 probability influence, with the next most relevant being bird’s claw, another severe schistosomal periportal fibrosis pattern that represents a blocked main portal vein, occluded vessels, and extensive fibrosis extending to the liver capsule (Niamey protocol pattern F) with 0.027 probability.

**Figure 1:**
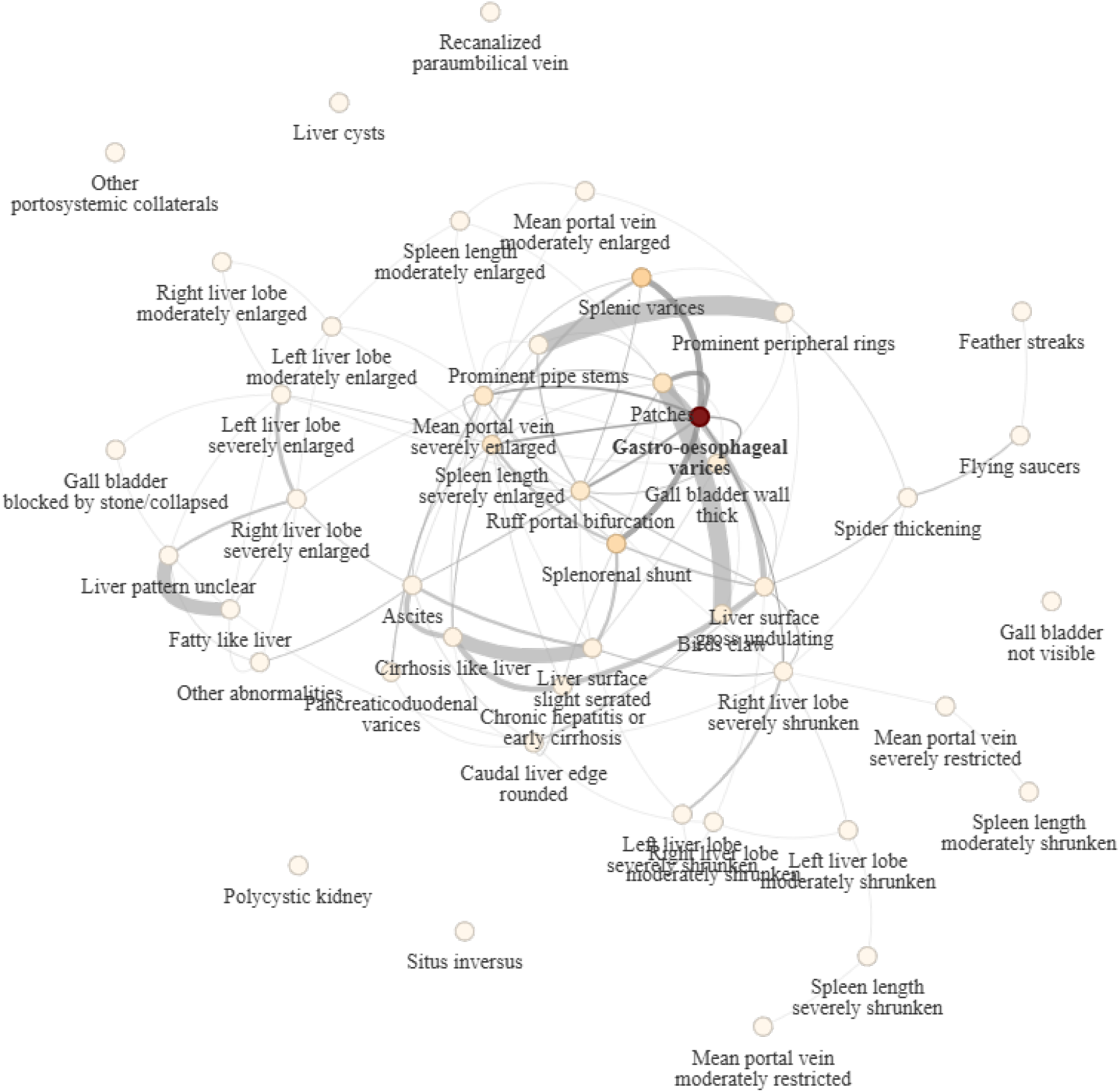
Influence probabilities on predicting gastro-oesophageal varices from models trained on other conditions. Probability on each node determines the proportion of influence from the model trained on the node condition on predicting gastro-oesophageal varices, darker colour implies larger influence, lighter colour implies more minor influence. Probabilities are obtained from the row corresponding to gastro-oesophageal varices in the graph convolution matrix *f*_*α*_(𝒢), *α* is chosen as the median of its posterior (see Fig. S5). The exact probabilities of the conditions can be found in Supplementary Fig. S7.

For periportal fibrosis (Niamey protocol patterns C to F), all patterns had self-influence probabilities greater than 0.4 (Supplementary Fig. S8 - S12). The patterns were highly inter-dependent and featured in each other’s dependencies. The D grade, being the intermediate grade between assumed fibrosis progression from C1/C2 to E was in the top five for all three with influence probabilities from 0.023 - 0.038 and 0.030, respectively. C1 and C2 are two different transections of the liver to capture similar levels of vessel-related fibrosis, and expectedly exhibited strong dependence (0.275 - 0.278 probabilities). The thickening and presentation of many second order branches (spider thickening fibrosis, Niamey protocol pattern B2, a non-periportal fibrosis associated liver pattern), also appeared in the top five for both (0.018 - 0.032 probabilities). The most severe grades of schistosomal periportal fibrosis, which were anatomically nested (Niamey protocol patterns E and F) also were strongly connected (> 0.175 probabilities), and strikingly both were highly dependent on a gross undulating liver surface (0.041 - 0.069 probabilities).

### Multimorbidity prediction

Predictive performances were broken down for the individual conditions presented in Fig. 2, with an average AUC across 45 conditions over 10 splits of 0.721 *±* 0.011 from the multitask model, compared to 0.636 *±* 0.007 from using the conventional 45 frequentist single-output models. The multitask model produced higher AUCs on 41 out of 45 conditions, with significant difference on 15 based on *t*-tests. Among the 15, nine were rated severe, including gastro-oesophageal varices with an AUC of 0.904 by the multitask model, and only 0.574 from the single-output model. Of the four conditions the multitask model underperformed on, all differences were insignificant.

**Figure 2:**
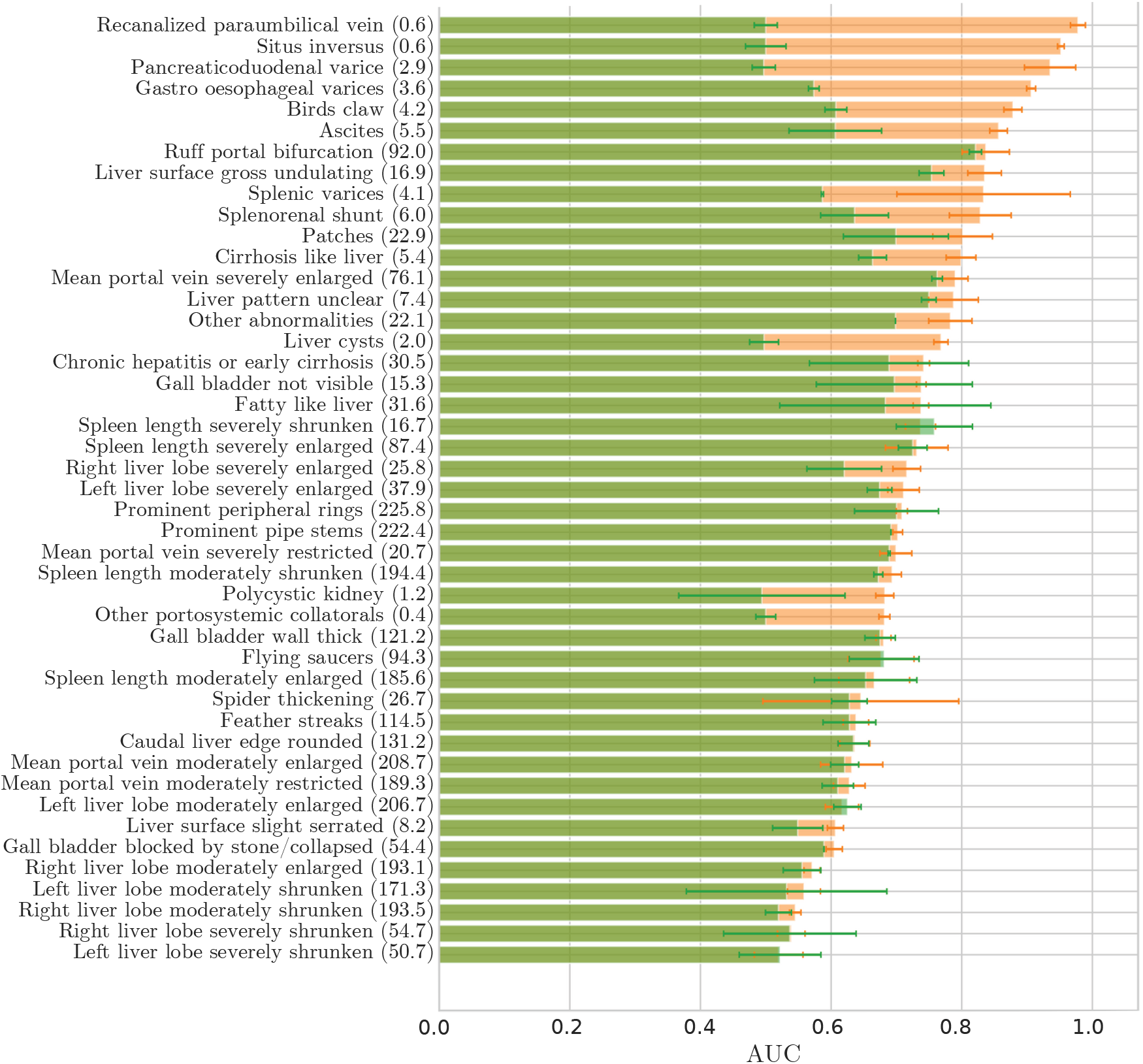
Conditions prediction AUCs. The AUC of each condition is averaged over 10 random training-testing splits. Average number of positive outcomes of each condition in the test set is shown in brackets. Average test AUC from multitask model (orange): 0.721 *±* 0.011, from 45 single-output models (green): 0.636 *±* 0.007.

When comparing different frameworks for predicting multiple outcomes, using multiple single-output models remained the worst performing approach by a significant margin, with even the Bayesian multi-output model out-performing with AUC of 0.665 *±* 0.019 despite forcing the same covariates for all outcomes. All models under our multitask framework performed better than the single and multi-output models, but using only the inclusion probabilities improved by a larger margin than using only the graph for condition inter-dependencies, with 0.718 *±* 0.012 and 0.694 *±* 0.011 respectively. The best performing model, producing 0.721 *±* 0.011 and the focus of the results, was with both multitask learning elements. These results also are visualized in Supplementary Fig. S4.

## Discussion

Schistosomiasis-related morbidities are poorly understood with no clinical or WHO guidelines for case management [14]. Schistosomiasis is not a single disease but a complex set of conditions (multimorbidity) arising partially due to past schistosome infection but also a wide range of diverse, often interacting causes. We studied a comprehensive list of risk factors and condition inter-dependencies for hepatosplenic multimorbidity. 3155 individuals aged 5-91 were clinically assessed for 45 liver and spleen conditions using point-of-care ultrasound within a community-based cohort (SchistoTrack [21]) in rural Uganda. Bayesian multitask learning models were developed for identifying risk factors across the 45 conditions where we accounted for condition influences using a graph structure and shared influence probabilities. Here we show that hepatosplenic multimorbidity is common where schistosomiasis is endemic and is associated to diverse biomedical, social, and spatial risk factors as well as inter-dependencies between biologically inter-related conditions.

Older age was a strong consistent positive predictor of a large number of diverse individual hepatosplenic conditions, the group of liver patterns representative of periportal fibrosis, and overall hepatosplenic multimorbidity. The finding of older age and multimorbidity is consistent with studies of widely different diseases, e.g. cardiovascular diseases [6]. However, our study now demonstrates the importance of age even in the context of chronic infections, suggesting that age is not simply an indicator of multimorbidity in LMICs due to the conventionally proposed effects of increased frailty with natural aging [5]. Our study population was relatively young compared to the life expectancy in high-income countries, with 87% of individuals less than 50 years. However, the positive association still reflects the association of aging and the longer time period available for acquiring multiple conditions as shown for general multimorbidity patterns [2, 16, 17, 22, 23]. Given the high endemicity of *S. mansoni, Plasmodium falciparum*, HBV, and HIV, the chronic or repeated exposure to these pathogens over the life course may also have been represented through age despite our study only measuring current infections. Future work, such as that ongoing in SchistoTrack [21], is needed to study the rate of multimorbidity accumulation over age that is attributable to chronic pathogen exposure, especially given the widespread availability of treatment and interventions for common pathogens contributing to individuals living longer with infectious diseases.

Higher Hb concentration was significantly associated with lower likelihoods of 11 hepatosplenic conditions. Most of these conditions encompassed characteristics of severe schistosomal morbidity related to portal hypertension including gastro-oesophageal varices, but also splenorenal shunts, ascites, and splenic enlargement. The associations were robust to sub-group analyses in adults only. Hb concentration may be relevant for hepatosplenic multimorbidity due to individuals losing blood from burst gastro-oesophageal varices [15]. Alternatively, the association of Hb with hypersplenism suggests splenic dysfunction and immune impairment resulting in excessive red blood cell sequestration [24]. Chronic infection with *S. mansoni* also contributes to low Hb concentration [25]. Hb and hypersplenism were associated independently of malaria, which had no association with any splenic enlargement in adults. It is unlikely the association of Hb and hypersplenism is due to sickle cell disease where more commonly hypersplenism is observed in children who are treated with a splenectomy before reaching adulthood [26]. Importantly, being a fisherman, which was an indicator of past schistosome exposure [27], was positively associated with splenic enlargement. Hb concentrations are already used as a sign of patient stability in emergency care and general screening in routine primary healthcare practice. Hb is a broad indicator that can encompass many risk factors from genetic disorders, infections, poor nutrition, immune disorders, to recent traumatic blood loss among other causes [28]. Our findings reveal that Hb concentrations are informative for screening for severe schistosomal morbidities. Given its routine measurement in clinical care and low cost, Hb concentration should be investigated in future prospective studies as an early indicator used for triage and risk classification of complex hepatosplenic multimorbidity. As a start, we found differences in WHO anaemia categories [28] related to severe conditions, but additional research is needed to define Hb thresholds to match levels of multimorbidity severity, as tied to progression and prognosis. Once known, national clinical guidelines in countries where schistosomiasis is endemic should consider Hb thresholds in case management protocols for portal hypertension. By contrast, current alcohol use was not associated with any hepatosplenic conditions despite being a regularly assessed risk factor for portal hypertension by clinicians [29]. Additional studies beyond current alcohol use are needed that consider the history of consumption and quantity.

Easily observable participant characteristics related to demographics, socioeconomic status, and location also were relevant risk factors for hepatosplenic multimorbidity. Males were more likely to have more hepatosplenic conditions, in addition to characteristics of being a fisherman and proximity to open freshwater sites. Men are more likely to engage in activities involving water contact, such as fishing, that determine the history of exposure to schistosome infection [27, 30], which is of greater relevance than current infection for periportal fibrosis [10, 13]. Living in either Buliisa and Pakwach districts had positive effects on overall multimorbidity compared to Mayuge, with the highest likelihood related to Pakwach. This result suggests there is spatial clustering in multimorbidity that needs to be accounted for in future studies. The district effect may also capture high-level risk factors such as the access to and quality of healthcare. Pakwach was the only district in our study that did not have a hospital, instead operating with only one dilapidated and understaffed Health Centre IV, and was often low on emergency medicine kits (saline and other supplies) and blood for transfusion that would be needed to respond to upper gastrointestinal bleeding from burst gastro-oesophageal varices. Pakwach Health Centre IV also lacks essential sonography expertise and ultrasound equipment to non-invasively diagnose gastro-oesophageal varices. The lack of supplies coupled with inadequate triage and investigation due to lack of knowledge of risk factors by health workers may result in premature mortality of patients. The geographical disparities highlighted in this study identify a need for recruitment of additional health staff, education on schistosomiasis-related morbidities, sonography training, and provision of supplies and equipment in the areas with the highest burden of hepatosplenic multimorbidity.

We identified risk factors for gastro-oesophageal varices, which indicate severe portal hypertension and pose several challenges for early detection in areas with limited diagnostic resources including a lack of endoscopy units [31, 32]. Only older age and lower Hb concentration were positively associated to the likelihood of having gastro-oesophageal varices. Although insignificant, there were non-zero positive effects from HIV, HBV, being a fisherman, and being male, with the largest effect size from HIV. Notably, HIV, being a fisherman, and being male were significantly positively associated with schistosomal periportal fibrosis (Niamey protocol patterns C-F) that precedes portal hypertension complications such as gastro-oesophageal varices. These risk factors have been shown elsewhere to be relevant either due to interacting biological causes (HIV) or representing a history of schistosome exposure (fisherman and gender) [10]. Additional prospective studies are needed to confirm the mechanistic relationship with schistosome-HIV co-infections, as well as to investigate morbidities specific to HIV but not to schistosomiasis.

Importantly, we discovered several clinical risk factors from the condition dependencies found in the multitask model for gastro-oesophageal varices. There was a higher probability of an individual having gastro-oesophageal varices when only occluded vessels and fibrosis blocking the main portal vein (Niamey protocol pattern E, patches), or fibrosis at the point of portal bifurcation were observed (Niamey protocol pattern D, ruffing). There were no associations with current schistosome infection. These findings show that only severe schistosomal periportal fibrosis patterns, which require a long history of schistosome exposure or interactions with co-infections, increase the likelihood of gastro-oesophageal varices. Notably, there were conferred risks from hypersplenism, a severely enlarged main portal vein, and various portosystemic collaterals including splenic varices and splenorenal shunts. Our study provides a shortlist of conditions and portal hypertension complications that should be prioritized as danger signs for future clinical guidelines seeking to preemptively identify patients at risk of developing gastro-oesophageal varices and manage patients with existing varices.

Our study makes several major methodological contributions for modelling multimorbidity. Standard logistic regressions struggle with class imbalance, and we observed low AUCs for these models on a number of sparse outcomes. However, class imbalance is inherent to hepatosplenic conditions. Gastro-oesophageal varices, for example, are an end-stage complication of portal hypertension and often with survivor bias (given associated high mortality rates). This problem was mitigated in the multitask setting as each model was able to learn from the models for other potentially more common conditions. This importance was demonstrated by the relevance of the condition dependencies over covariates for infrequent outcomes such as gastro-oesophageal varices. Multitask learning exposes each condition to similarly behaving and biologically-related conditions to maximize available information. Most conditions, including gastro-oesophageal varices, saw benefits from learning of risk factors from other related conditions as evidenced by significantly improved AUCs. The individual AUCs for each condition from multitask modelling were superior for 91% (41/45) of all conditions and significantly different for 15 conditions.

Our study goes beyond the conventional use of graphs for multimorbidity where condition dependencies are typically inferred only between directly connected nodes [33, 34]. The global graph convolution function accounted for edge weights and graph distance, allowing the strength of relationships between nodes further away to also be quantified. Additionally, while previous work has utilized similar variable selection procedures through inclusion probabilities [33, 34], they have been treated as discrete binary variables, which are less flexible than continuous inclusion probabilities that could encompass both good and uninformative associations between each covariate and the range of outcomes.

Our study has some limitations. The current framework does not account for task importance and treats all conditions equally irrespective of severity. Clinically, the inferred dependencies between conditions suggest biological relatedness, but they do not imply causality, and therefore cannot be used to explain pathogenesis. The identified risk factors are informative for clinical management, but require additional research to account for genetic risk and identifying immunological mechanisms. Although inferring in a fully Bayesian manner greatly improved the flexibility of the parameters, quantifying associations with heavily skewed posterior distributions such as those with HIV and HBV was more difficult. Meanwhile, the graph was utilized to quantify inter-task strengths for finding related conditions as risk factors and allowed us to observe the influence of conditions without including them as input covariates. However, the chosen graph function, despite providing improvements to the prediction, was not as important as the risk factors. Future work should investigate more flexible deployments of the graph to incorporate properties of controlling condition importance and structural learning.

Hepatosplenic multimorbidity is a major global health challenge due to the wide range of biological, social, and spatial causes, high prevalence in resource-constrained settings, and the complexity of identifying condition dependencies to inform clinical guidelines. This study validates the usefulness of Bayesian machine learning in addressing these challenges. We have demonstrated the performance of Bayesian multitask learning for predicting hepatosplenic multimorbidity in schistosomiasis-endemic areas, while the framework has wide generalisability to other multimorbidities. We discovered key risk factors and condition dependencies of 45 schistosomiasis-related hepatosplenic conditions that could be used to develop clinical and WHO guidelines for morbidity identification and case management in endemic countries.

## Methods

### Participant sampling

This study was conducted within the SchistoTrack cohort [35] during the first annual follow-up between 17 January and 16 February 2023. Participants were selected from 1952 randomly sampled households from 52 villages across Buliisa, Pakwach, and Mayuge Districts of Uganda; 38 of the villages were sampled in the baseline of 2022 [10]. One child aged 5-17 years and one adult aged 18-91 were selected by the household head or spouse and invited for clinical assessments. 3224 individuals were examined with point-of-care ultrasound in 2023. There were 3186 individuals out of 3224 with non-missing ultrasound data, and were featured in learning the graph [9] utilized for this study. All participants had household demographic covariates data, but 3155 had non-missing data from blood and stool assessments and were considered for analysis in this work. A breakdown of missing data can be found in the participants flow diagram in Supplementary Fig. S1.

### Hepatosplenic outcomes

The outcomes were hepatosplenic conditions obtained by point-of-care ultrasound. Philips Lumify C5-2 curved linear array transducers were used with the Philips Lumify Ultrasound Application v3.0 on Lenovo 8505-F tablets with Android 9 Pie. Lossless DICOM images and videos were saved for quality assurance [10]. We assessed a total of 45 hepatosplenic conditions listed in Table 1. The conditions were measured as indicators as described in [9], including focal and diffuse liver fibrosis patterns following the WHO Niamey protocol, liver surface irregularities, caudal liver edge assessments, fatty and cirrhotic livers, liver and spleen organometry, portal vein dilation or restriction, portosystemic collaterals, ascites, gall bladder obstruction, among others. For the left and right liver lobes, spleen, and portal vein diameter, we considered abnormality as one or two standard deviations above or below an internal healthy reference population from the same year split by height, as described in [9]. Importantly, all conditions were assigned a severity rating in relation to schistosomal portal hypertension as shown in the final column, these were determined from expert opinion by a clinical epidemiologist (GFC), sonographer (VA), and gastroenterologist (CKO) as detailed in [9].

### Participant covariates

Participants covariates were collected through a combination of household surveys, and diagnoses by laboratory technicians and nurses. These are listed in Table 3, detailed definitions can be found in [10, 21]. Clinical measurements included *S. mansoni* status as measured by microscopy and eggs per gram, malaria rapid diagnostic test outcomes, HBV rapid diagnostic test outcomes, self-reported HIV, and point-of-care Hb concentration. Although not a model covariate, anaemia status was investigated by constructing indicators based on WHO definitions where thresholds for mild, moderate, and severe anaemia considered age, gender, and pregnancy [36]. Socio-demographic information included age, gender, belonging to the majority tribe of their district, belonging to the majority religion of their district, years of educational attainment, occupations (farmer, fisherman, and fishmonger, with unemployed and other occupations as reference). Home quality score was calculated based on the quality of materials used to build the house. Individuals belonged to a household with social status if any adult member held or previously held a position in the local council, village health team, religious or clan leadership, or influential beach management committees. Additional household variables included number of individuals in the household, number of years household lived in the village, if they owned the home they lived in, number of rooms in the household, current alcohol use based on self-reported consumption within the year preceding recruitment, improved water source defined by if they obtained water from any of protected well or spring, borehole, village tap, and rainwater, number of water activities, and the year of recruitment (2022 or 2023, with the baseline recruitment year of 2022 as reference). Finally, locational variables measured the distance (in kilometres) to nearest freshwater site, and government health centre, and the respective districts (Buliisa, Pakwach, with Mayuge as reference). The data was standardized pre-modelling to make priors easier to choose as detailed in later sections.

**Table 3:** List of covariates. Reference categories of categorical variables are italicized. Summaries are calculated from a total of 3155 participants. When log transforms are taken, summaries of the raw values are also provided but are not used for modelling.

| Covariate | Category | Summary Type | Summary |
| --- | --- | --- | --- |
| *S. mansoni* eggs per gram (log(+1)) |  | Mean (SD) | 1.40 (2.19) |
| *S. mansoni* eggs per gram (raw) |  | Mean (SD) | 85.25 (404.28) |
| Malaria rapid diagnostic test |  | Count (%) | 1469 (46.56) |
| HBV |  | Count (%) | 179 (5.67) |
| HIV (self reported) |  | Count (%) | 55 (1.74) |
| Hemoglobin concentration (log) |  | Mean (SD) | 2.51 (0.22) |
| Hemoglobin concentration (raw, g/dL) |  | Mean (SD) | 12.62 (2.53) |
| Age |  | Mean (SD) | 25.93 (18.39) |
| Gender | *Male* | Count (%) | 1457 (46.18) |
|  | Female | Count (%) | 1698 (53.82) |
| Majority tribe |  | Count (%) | 2404 (76.20) |
| Majority religion |  | Count (%) | 2420 (76.70) |
| Years in education |  | Mean (SD) | 3.25 (2.78) |
| Occupation | *Other/None* | Count (%) | 2296 (72.77) |
|  | Farmer | Count (%) | 540 (17.12) |
|  | Fisherman | Count (%) | 219 (6.94) |
|  | Fishmonger | Count (%) | 100 (3.17) |
| Home quality score |  | Mean (SD) | 5.60 (3.41) |
| Household social status |  | Count (%) | 385 (12.20) |
| Number of individuals in household |  | Mean (SD) | 3.65 (1.42) |
| Years household lived in village |  | Mean (SD) | 20.10 (15.85) |
| Home owned |  | Count (%) | 2759 (87.45) |
| Number of rooms |  | Mean (SD) | 2.14 (1.19) |
| Current alcohol use |  | Count (%) | 306 (9.70) |
| Improved drinking water source |  | Count (%) | 1748 (55.40) |
| Number of water activities |  | Mean (SD) | 1.76 (0.97) |
| Year of recruitment | *2022* | Count (%) | 2207 (69.95) |
|  | 2023 | Count (%) | 948 (30.05) |
| Min. dist. (km) to water site |  | Mean (SD) | 0.52 (0.44) |
| Min. dist. (km) to health centre |  | Mean (SD) | 4.11 (3.31) |
| District | *Mayuge* | Count (%) | 754 (23.90) |
|  | Buliisa | Count (%) | 1006 (31.89) |
|  | Pakwach | Count (%) | 1395 (44.22) |

### Model specification

We adopted a Bayesian multitask model for predicting the 45 hepatosplenic conditions diagnosed in this study. Multitask models have often been presented as neural networks (e.g. [37–40]), but we proposed an architecture and inference framework that was arguably more general and suitable for clinical studies. Importantly, existing work in Bayesian multitask regression [41–43] lacked our level of interpretability. The model was inferred in a Bayesian manner such that the possible relationships between covariates and outcomes were incorporated more comprehensively as distributions [44], while we were able to maintain interpretations of significance and strength of associations. A Bayesian procedure was implemented to also improve the handling of imbalanced outcomes, as was the case for many severe conditions. Having such uncertainty in the model had been shown to be robust against class imbalance [44], mitigating the need for resampling methods such as SMOTE [45]. Importantly, we designed a new model architecture following the properties of multitasking, introducing ways for parameters to be exposed to multiple outcomes. All factors incorporated led to a final model with far superior predictive performance. Each step of model interpretation and utilization is summarized in Fig. 3.

**Figure 3:**
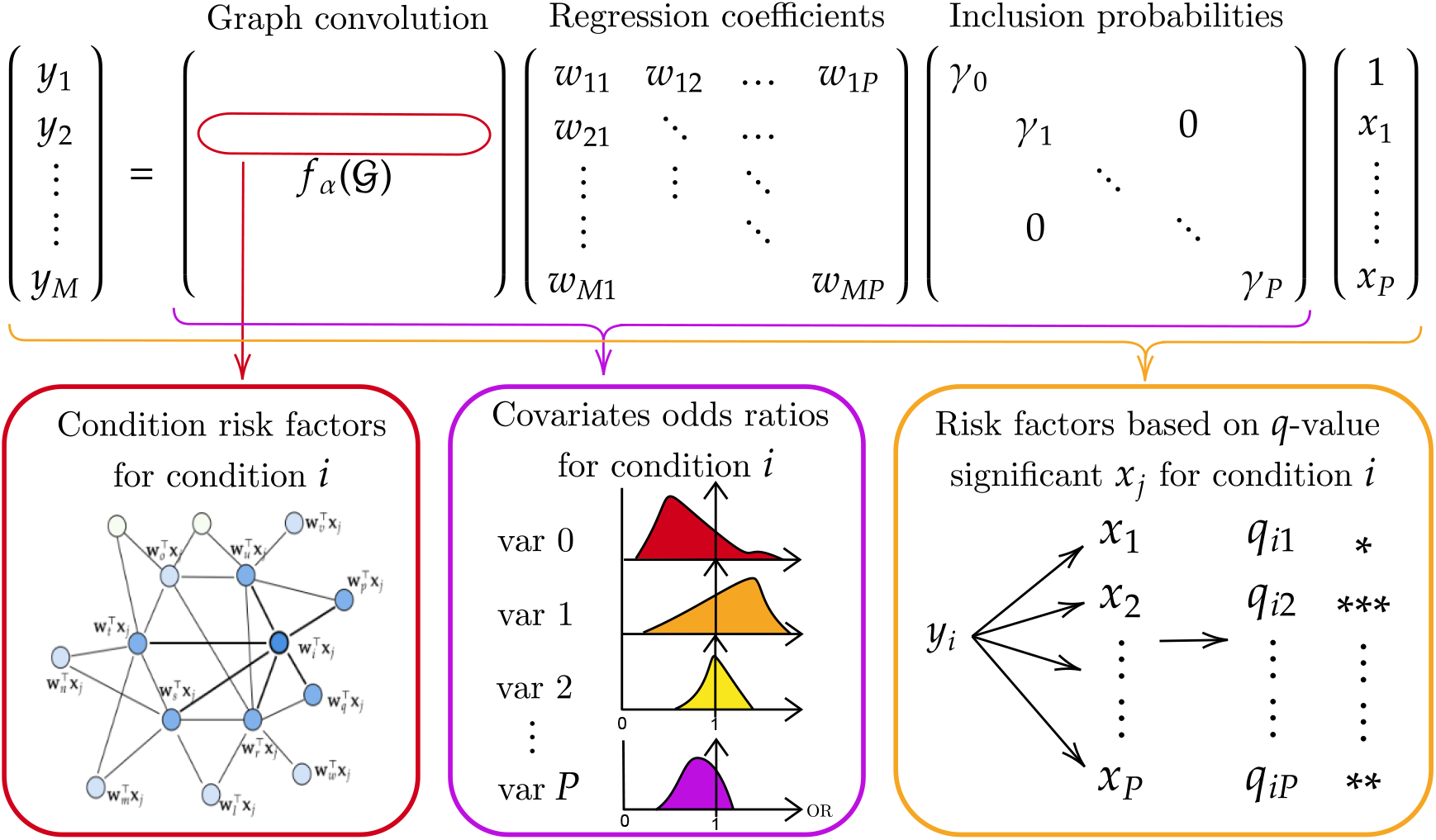
Pipeline of study.

Models were developed to predict *M* = 45 possible hepatosplenic conditions within an individual, participant data were indexed by *i* = 1, …, *N*. The outcome conditions were represented by indicator vectors **y**_*i*_ = (*y*_*i*1_, …, *y*_*iM*_)^⊤^, such that *y*_*ij*_ = 1 if person *i* has condition *j*, and 0 otherwise. Each participant was represented by (**x**_*i*_, **y**_*i*_), where we predicted the set of conditions **y**_*i*_ given the individual covariates **x**_*i*_. We accounted for dependencies between the conditions through a thresholded multimorbidity graph 𝒢 representing positive inter-relationships between the 45 hepatosplenic conditions; the graph used was learned and validated following the pipeline from previous study [9].

To progress towards multitask modelling, we started with single outcome logistic regression defined as

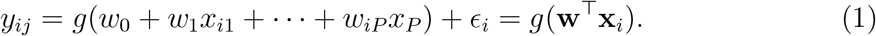

In the case when *M* outcomes (*M >* 1) existed for analysis, we built *M* models for person *i* by

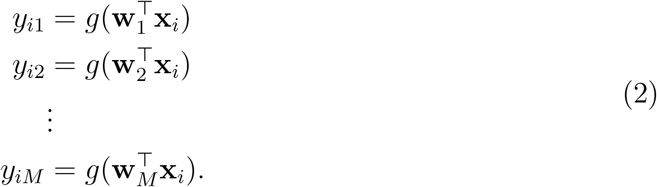

For more compact notation, we combined the regression weights/coefficients (**w**_*i*_) as

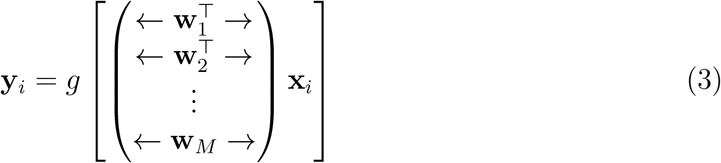

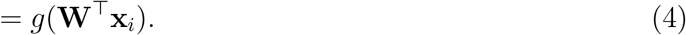

Model (4) was the basis of multi-output modelling [46], where it was important to note that *y*_*ij*_ ∈ **y**_*i*_ continued to be predicted by 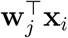 only.

We extended model (4) to a multitask framework by proposing the introduction of two new terms:

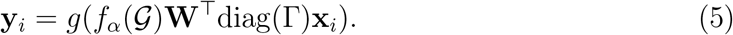

*f*_*α*_ is a function defined from the graph, and Γ = (*γ*_0_, …, *γ*_*P*_)^⊤^ is a vector of probabilities for the *P* possible covariates. The roles of the two terms were to introduce multitask model structures as explained in the following sections.

### Multitask graph convolution

We introduced the graphical function *f*_*α*_(𝒢) ∈ ℝ^*M ×M*^ that convolved the models on each outcome. The aim was to replicate the known inter-relatedness of biological interactions of hepatosplenic conditions by introducing inherent relational inter-dependence between the models on each condition. Without this term, relations between the models would not be incorporated in the model. We chose the graphical function from signal processing [47, 48] to act as filtering functions to smooth node data based on the graph

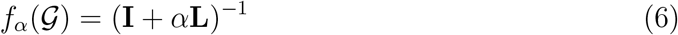

where **L** was the regularized graph Laplacian defined by

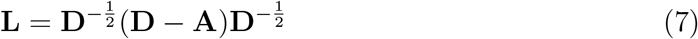

for adjacency matrix **A** and diagonal degree matrix **D** based on the graph, and *α* is an additional positive parameter controlling the influence of the graph.

By multiplying the multi-output model with *f*_*α*_(𝒢) ∈ ℝ^*M×M*^, each outcome would be predicted by not only the corresponding model, but also the models on proximate nodes based on the graph structure. The choice of *f*_*α*_ allowed the construction of continuous weightings that accounted for both edge weights and graph distances. The weights assigned to the nodes were further controlled by the parameter *α* and optimized as part of the model inference. The *i*th rows of *f*_*α*_(𝒢) provided the weighting across the graph with the *i*th node as centre, ensuring model *i* primarily focused on condition *i* but drawing influence from models on proximate nodes. The size of the weights exhibited similar scale to probabilities, and were therefore normalized to one so that they could be interpreted as such. Condition *i* would be predicted by a weighted average of models, with the weighting on proximate nodes indicating the additional likelihood of developing condition *i* contributed from the other models. The value on node *i* itself would be interpreted as self-influence, reflecting solely the model developed for that condition and therefore the influence of the covariates on the singular condition. Isolated nodes in the graph represented unrelated conditions and would be treated as independent heterogeneous tasks. The graph filtering function (6) provided weightings for all nodes, incorporating more information than existing studies on multimorbidity networks which restricted analysis to 1-hop neighbourhoods. As the conditions came in groups based on different diagnoses of sonography (see first column of Table 1 for grouping), some conditions naturally co-occurred, therefore, our analysis also differentiated the two types of influences, with conditions from different groups classified as risk factors, while same group connections were interpreted as co-occurring conditions.

### Multitask covariate learning

The term diag(Γ) ∈ ℝ^(*P* +1)*×*(*P* +1)^ performed the role of multitask covariate learning, with each *γ*_*i*_ acting as global inclusion probabilities of the covariates across all models. When combined with the covariates, diag(Γ)**x**_*i*_ = (*γ*_0_, *γ*_1_*x*_*i*1_, …, *γ*_*P*_ *x*_*iP*_)^⊤^ (assuming the first term in the covariate vector was the intercept). Each *γ*_*i*_ was inferred to represent the utility of covariate *i* for all outcomes as a joint distribution, allowing for covariate information sharing between models. For categorical variables, the categories were encoded as indicators, but we enforced the same inclusion probability across all indicators of the same variable. For instance, if *x*_*ic*1_, *x*_*ic*2_, and *x*_*ic*3_ were possible outcomes of *x*_*c*_, then diag(Γ)**x**_*i*_ = (*γ*_0_, …, *γ*_*c*_*x*_*ic*1_, *γ*_*c*_*x*_*ic*2_, *γ*_*c*_*x*_*ic*3_, …)^⊤^. Doing this provided a measure of relevance of *x*_*c*_ as a whole instead of each indicator independently.

While the inclusion probabilities acted as global multipliers shared across all models, inferring them in a Bayesian manner expanded their function as a parameterization of all the possible associations between the covariates against multiple outcomes. Both favorable (high probabilities) and uninformative (low probabilities) associations would be reflected in a single posterior distribution, which were fully incorporated for prediction.

The role of inclusion probabilities could also be interpreted in the context of Bayesian variable selection, drawing similarities to the spike-and-slab prior when coupled with the regression coefficients. Each *γ*_*i*_ could potentially induce shrinkage effects on the covariates, and as a result, covariates were standardized before model fitting to ensure equal, comparable shrinkage, as consistently practiced with common Bayesian variable selection priors [49].

### Prior selection

The multitask model took the following form:

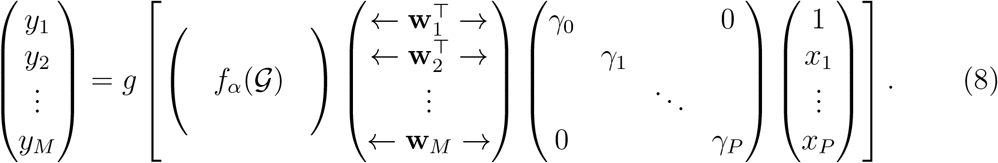

We henceforth refer to *f*_*α*_(𝒢) as the graph convolution, **W** as the regression coefficient matrix, and Γ as the inclusion probabilities.

The model was inferred in a Bayesian manner with all parameters given priors, and predictions were found by marginalization [44, 50]. First, inclusion probabilities were assigned priors with support between [0, 1]:

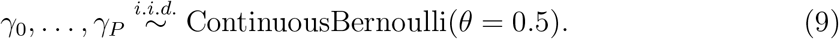

The continuous Bernoulli was chosen due to its similarity to the binary counterpart when *θ* was close to 0 or 1 [51, 52], making each *γ*_*i*_ easier to interpret as inclusion/exclusion.

As regression weights *w*_*ij*_ could be positive or negative, we used a common Gaussian distributions with varied variance

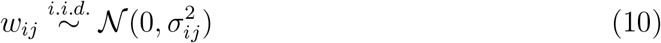

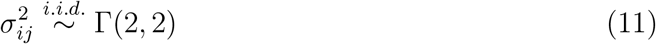

such that positive priors were assigned to the variance of each Gaussian distribution. This setup allowed the regression weights to have non-fixed variance to adapt to the uncertainty of the covariates. The Gamma distribution was chosen because of its non-negativeness and flexible shape, its hyperparameters were chosen to have mode 1, made appropriate by the normalization of the covariates.

Concerning the graph convolution with non-negative parameter *α*, we assigned the same positive prior as previously described to provide a suitable starting point during inference

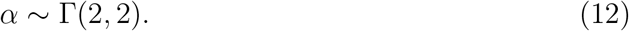

### Bayesian inference

Bayesian machine learning is characterized by the inference procedure involving integrating over the uncertainty over all parameters. This was the key advantage of the Bayesian approach, as predictions were based not on single point estimates of the parameters, but Bayesian averaged over all possible parameter configurations weighted by their posteriors. Here, we detail the steps to carry out Bayesian inference on the model.

Model parameters were inferred using Markov Chain (Hamiltonian) Monte Carlo (MCMC) [53, 54] using the NUTS kernel from probabilistic programming library Pyro [55]. Let Θ = {Γ, **W**, *σ*_2_, *α*} represent all parameters of the multitask model *f* (**x**_*i*_) = *g*(*f*_*α*_(𝒢)**W**^⊤^diag(Γ)**x**_*i*_), and data 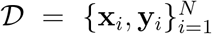. We were interested in obtaining the posterior distribution

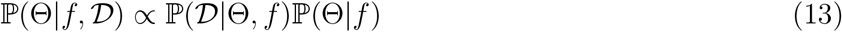

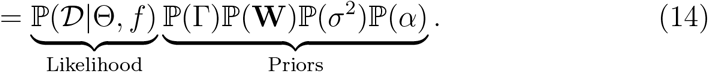

The priors above were defined by (9), (10), (11), and (12). For binary classification, the likelihood was the Bernoulli density:

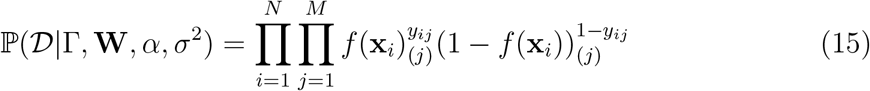

over *N* participants and *M* conditions, where *f* (**x**_*i*_)_(*j*)_ denoted the *j*th element of *f* (**x**_*i*_) ∈ ℝ^*M*^. We assumed conditional independence of the outcomes in the likelihood, but statistical dependence was induced through the multitask parameters capturing interactions among outcomes. This setup allowed the likelihood to reflect outcome dependencies while retaining a computationally tractable likelihood. This was the optimal choice compared to multivariate logistic [56] or probit [57] likelihoods, which were intractable with the number of outcomes, and the dependence specification was through random effects as covariance structures instead of fixed effect of the multitask parameters.

Predictions on novel inputs **x**_*_ were obtained by computing

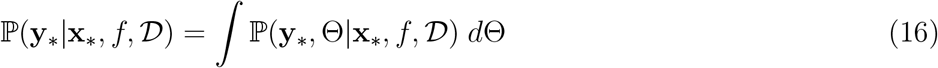

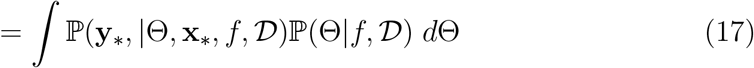

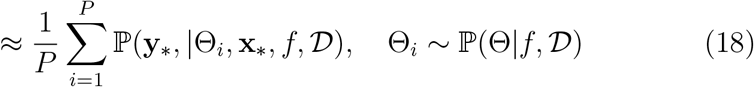

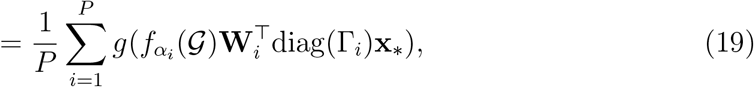

where each set of parameters 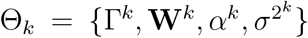 were samples of the posterior obtained by MCMC. Each sample represented a configuration of the model, and they corresponded to the highest posterior density and optimal parameters for model fit. As shown by equation (19), predictions were made by Bayesian averaging *P* models instead of one single model, this was the signature of Bayesian machine learning that led to far greater flexibility and predictive capacity.

### Covariate significance

Risk factors were obtained from the covariates that exhibited significant associations with the outcome conditions. The likelihood ratio test was used here, as while significance could be determined by examining credible intervals, posterior distributions were skewed and would lead to different conclusions depending on which credible interval to use. Using credible intervals alone also lacked *p*-values needed for false discovery rate correction as explained later.

Nested model log-likelihood ratios were well-known to follow the 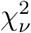 distribution, here, the degree of freedom was dependent on the number of outcomes. The likelihood (15) represented the full model for all outcomes, but for individual conditions, the partial log-likelihoods was computed by fixing *j* in (15) to the index of the covariate of interest

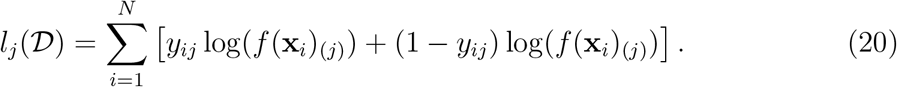

Let *l*_*j*_(𝒟_−*k*_) represent the log-likelihood evaluated with covariate *k* excluded, the test for covariate *k* against outcome *j* was computed through the likelihood ratio

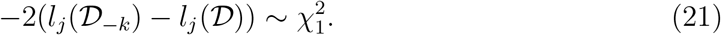

More importantly, we were able to measure covariate significance specific to groups of conditions by combining their log-likelihoods. This was useful for examining any multimorbid complications consisting of specific sets of conditions, such as the multiple liver fibrosis patterns indicative of periportal fibrosis (Niamey protocol patterns C - F). The likelihood ratio test became

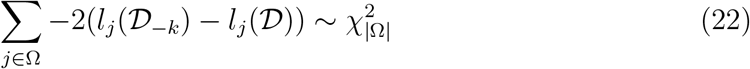

where Ω represented the set of conditions of interest, and the degree of freedom became the size of Ω because covariate *k* appeared once in the model on each condition. Summing over all 45 conditions would lead to a measure for overall hepatosplenic multimorbidity.

The large number of tests computed here would lead to high false discovery rate, thus, we controlled this by converting the *p*-values to *q*-values [58, 59] to determine the significance. The categories in occupation and district were treated as separate variables as they consisted of individual coefficients, thereby taking up a degree of freedom each. Thus, there was a total of 28 covariates to test, and we adjusted for the 28 *×* 45 = 1260 tests for all individual conditions, and 28 tests for grouped multimorbidity.

### Risk factor association

We interpreted the strength and direction of risk factor association by converting model posteriors to odds ratios. Each element of the model - graph convolution, regression coefficients, and inclusion probabilities - had individual posteriors, but we used the joint distribution of the three combined to reflect the scale of the data. Odds ratios were outcome specific such that for covariate *j*, we take the entry [*f*_*α*_(𝒢)**W**^⊤^diag(Γ)]_*ij*_ to find the association with condition *i*. Division by the standard deviation of each covariate was applied such that the odds could be interpreted in the original units (undoing the data standardization prior to model inference).

More precisely, let *m*_*ij*_ represent the *ij*-th entry of [*f*_*α*_(𝒢)**W**^⊤^diag(Γ)], and 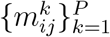 represent the MCMC samples, we examined the empirical distribution of the odds ratio by the points 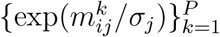 where *σ*_*j*_ was the standard deviation of covariate *j*. When considering grouped conditions, we combined the MCMC samples for each condition to provide the overall effect.

The posterior densities were generally skewed and asymmetric, thus, we reported the median when inspecting individual covariates, but used the mean for overall effects. Credible intervals to 95% were computed using the highest posterior density [60], which were better equipped to handle skewed distributions than equal-tailed intervals.

### Frameworks for predicting multi-outcome problems

Different frameworks were tested for handling multiple outcomes to compare against conventional approaches. As the most conventional approach, multiple frequentist single-output models were ran for each outcome with individual variable selections using Akaike information criterion (AIC). Consequently, each of the 45 frequentist models would consist of different covariates. For standard multi-output modelling, we used model (4) with a common set of variables, parameters were found through Bayesian inference as no variable selection existed for multi-outcome setting.

Multitask modelling induced inter-dependencies by introducing a small number of additional parameters, thus allowing the model to remain interpretable by maintaining regression model structures. We ran various versions of our multitask modelling. First, we tested multitask modelling via graph convolution, using model (8) without the covariate learning term, focusing on inter-task dependencies induced through the graph. Next, we ran multitask modelling via covariate learning, using model (8) without the graph convolution matrix, measuring the utility of the inclusion probabilities. The main model in this study, where results presentation focused on unless otherwise stated, incorporated both multitask learning elements via graph convolution and multitask covariate learning, taking the full form of model (8).

### Model prediction and evaluation

Prior analysis had been performed using 100% of the data, but to compare the predictive performance of multitask modelling against other frameworks, we now divided participants into training and testing sets of sizes 50% each (1593). The sets were randomly sampled, and we completed this ten times to obtain an overall evaluation of the predictions. This procedure was chosen instead of cross validation because some conditions occurred infrequently, and for these cases, random sampling would lead to more splits with at least one positive case appearing in the test set, allowing the model to be evaluated on infrequent conditions more often.

The training stage involved sampling from the posterior distributions ℙ (Θ |𝒟, *f*) using (Hamiltonian) MCMC, providing us with a set of posterior sample points for all parameters. For each split we ran 500 warm-up steps followed by 1000 posterior sampling steps. To make predictions given the test participants, the MCMC samples were used to compute the Monte Carlo estimator (19). Predictions were vectorial and binary, and evaluation was done for each outcome separately using the area under the receiver operating curve (AUC).

## Data Availability

The model implementation code is shared as supplementary material. Individual participant data for the predictions could not be shared due to their identifiable nature and associated ethics restrictions, data privacy considerations, and the ongoing nature of the SchistoTrack cohort.

## Acknowledgments

We are thankful for the involvement from our study participants, and the SchistoTrack teams especially the surveyors, nurses, sonographers, and laboratory technicians. We also like to thank the Uganda Ministry of Health, local district leaders, focal health workers, and village health teams. Special thanks also to the Oxford team for the fieldwork, data wrangling, everyday discussions, and feedback.

## Ethics approvals

Data collection and use were reviewed and approved by Oxford Tropical Research Ethics Committee (OxTREC 509-21), Vector Control Division Research Ethics Committee of the Uganda Ministry of Health (VCDREC146), and Uganda National Council for Science and Technology (UNCST HS 1664ES).

## Funding

This research was funded in whole, or in part, by the UKRI EPSRC [EP/X021793/1]. For the purpose of Open Access, the author has applied a CC BY public copyright licence to any Author Accepted Manuscript version arising from this submission. NDPH Pump Priming Fund, John Fell Fund, Robertson Foundation, UKRI EPSRC (EP/X021793/1) grants were awarded to GFC.

## Author contributions

Conceptualization: GFC and YCZ. Data curation: YCZ, BN, VA, JBO, NBK, CKO, GFC. Formal analysis: YCZ. Investigation, methodology, visualization: YCZ. Writing – original draft: YCZ and GFC. Validation: YCZ, GFC. Writing – review and editing: YCZ, GFC, BN, CKO, NBK, VA, JBO. Funding acquisition and supervision: GFC. Resources: GFC.

## Competing interest

The authors declare no conflicts of interest.

## Notes

### Competing Interest Statement

The authors have declared no competing interest.

