## Supplementary material for "Bayesian machine learning enables discovery of risk factors for hepatosplenic multimorbidity related to schistosomiasis"

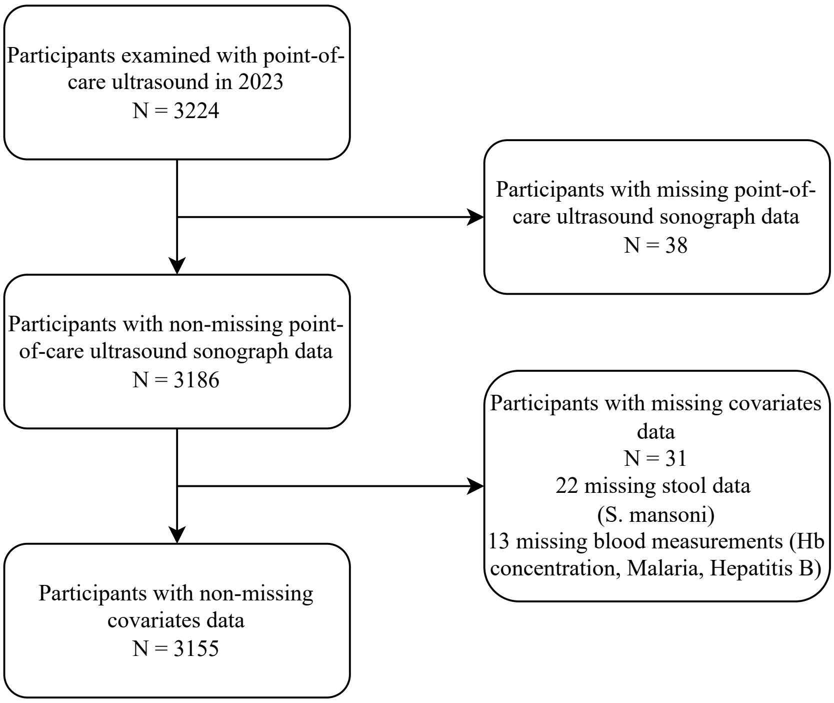


Figure S1: Participants flow diagram.

| Condition | Covariate | Median OR (95% CI) |
|---|---|---|
| Feather streaks | Pakwach | 1.43 (0.98, 2.05) |
| Prominent peripheral rings | Age | 1.04 (1.03, 1.05) |
| Prominent peripheral rings | Gender - female | 0.61 (0.48, 0.78) |
| Prominent peripheral rings | Buliisa | 1.48 (1.06, 2.05) |
| Prominent peripheral rings | Pakwach | 2.21 (1.55, 3.03) |
| Prominent pipe stems | Gender - female | 0.64 (0.50, 0.77) |
| Prominent pipe stems | Pakwach | 2.06 (1.52, 2.74) |
| Ruff portal bifurcation | Age | 1.08 (1.07, 1.09) |
| Ruff portal bifurcation | Gender - female | 0.53 (0.35, 0.80) |
| Patches | log(Hb concentration) | 0.20 (0.08, 0.58) |
| Patches | Gender - female | 0.60 (0.34, 1.00) |
| Chronic hepatitis or early cirrhosis | log(Hb concentration) | 0.15 (0.06, 0.38) |
| Chronic hepatitis or early cirrhosis | Pakwach | 2.59 (1.30, 5.15) |
| Left liver lobe moderately enlarged | Buliisa | 1.84 (1.30, 2.77) |
| Left liver lobe moderately enlarged | Pakwach | 1.95 (1.40, 2.88) |
| Left liver lobe severely enlarged | Age | 1.05 (1.03, 1.06) |
| Liver surface gross undulating | log(Hb concentration) | 0.21 (0.06, 0.51) |
| Liver surface gross undulating | Gender - female | 0.64 (0.31, 0.95) |
| Liver surface gross undulating | Pakwach | 3.94 (1.57, 11.66) |
| Caudal liver edge rounded | log(Hb concentration) | 0.25 (0.15, 0.47) |
| Caudal liver edge rounded | Buliisa | 1.94 (1.34, 3.01) |
| Mean portal vein severely enlarged | log(Hb concentration) | 0.17 (0.09, 0.34) |
| Mean portal vein severely enlarged | Age | 1.06 (1.05, 1.08) |
| Mean portal vein severely enlarged | Pakwach | 1.94 (1.35, 3.26) |
| Gastro-oesophageal varices | log(Hb concentration) | 0.07 (0.01, 0.37) |
| Splenorenal shunt | log(Hb concentration) | 0.08 (0.02, 0.41) |
| Ascites | log(Hb concentration) | 0.03 (0.01, 0.17) |
| Gall bladder not visible | Buliisa | 4.59 (1.46, 11.80) |
| Gall bladder wall thick | log(Hb concentration) | 0.54 (0.31, 0.86) |
| Gall bladder wall thick | Gender - female | 0.70 (0.52, 0.90) |
| Gall bladder wall thick | Pakwach | 1.52 (1.12, 2.22) |
| Spleen length severely enlarged | log(Hb concentration) | 0.07 (0.03, 0.13) |
| Spleen length severely enlarged | Pakwach | 2.63 (1.51, 4.16) |
| Spleen length severely shrunken | Age | 1.03 (1.01, 1.05) |
| Spleen length moderately enlarged | Age | 1.00 (1.00, 1.01) |
| Spleen length moderately enlarged | Pakwach | 1.78 (1.26, 2.45) |
| Spleen length moderately shrunken | Age | 1.00 (0.99, 1.01) |
| Spleen length moderately shrunken | Pakwach | 0.54 (0.39, 0.80) |

Table S1: List of significant relationships found between all conditions and covariates within the adults.
Adults are defined as 18 years or older in the year of study. Outcome conditions and their significant covariates are listed in the first two columns, significance is calculated to 5% based on corrected $q$-values.

| Condition | Covariate | Median OR (95% CI) |
|---|---|---|
| Chronic hepatitis or early cirrhosis | log(Hb concentration) | 0.05 (0.02, 0.46) |
| Right liver lobe moderately enlarged | log(Hb concentration) | 0.26 (0.20, 0.62) |
| Left liver lobe moderately enlarged | log(Hb concentration) | 0.16 (0.11, 0.44) |
| Left liver lobe severely enlarged | log(Hb concentration) | 0.05 (0.02, 0.38) |
| Caudal liver edge rounded | log(Hb concentration) | 0.16 (0.09, 0.43) |
| Caudal liver edge rounded | Min. dist. (km) to water site | 3.44 (1.80, 4.07) |
| Mean portal vein moderately enlarged | log(Hb concentration) | 0.23 (0.17, 0.66) |
| Gall bladder not visible | Buliisa | 2.77 (0.90, 5.45) |
| Gall bladder blocked by stone/collapsed | Gender - female | 0.63 (0.25, 0.95) |
| Spleen length severely enlarged | Age | 1.18 (1.05, 1.29) |
| Spleen length severely shrunken | log(Hb concentration) | 0.16 (0.02, 0.59) |
| Spleen length moderately enlarged | Malaria | 3.26 (2.83, 6.16) |
| Spleen length moderately enlarged | log(Hb concentration) | 0.15 (0.09, 0.34) |
| Spleen length moderately shrunken | Malaria | 0.16 (0.14, 0.25) |
| Spleen length moderately shrunken | Age | 0.83 (0.80, 0.91) |

Table S2: List of significant relationships found between all conditions and covariates within children.
Children are defined as 5-17 years old in the year of study. Outcome conditions and their significant covariates are listed in the first two columns, significance is calculated to 5% based on corrected $q$-values.

| Covariate | GOV Median OR (95% CI) | GOV $q$ | PPF Mean OR | PPF $q$ | Overall Mean OR | Overall $q$ |
|---|---|---|---|---|---|---|
| *S. mansoni* log(epg+1) | 0.94 (0.74, 1.04) | 0.78 | 0.97 | 0.50 | 0.99 | 0.11 |
| Malaria | 0.92 (0.52, 1.30) | 1.00 | 0.87 | 0.50 | 0.95 | <0.01 |
| HBV | 1.40 (0.81, 3.72) | 1.00 | 1.21 | 0.78 | 1.06 | 1.00 |
| HIV | 2.52 (0.97, 10.77) | 0.61 | 1.77 | 0.04 | 1.27 | 1.00 |
| log(Hb concentration) | 0.07 (0.01, 0.37) | 0.01 | 0.66 | 0.45 | 0.55 | <0.01 |
| Age | 1.04 (1.02, 1.08) | 0.01 | 1.04 | <0.01 | 1.02 | <0.01 |
| Gender - female | 0.67 (0.26, 1.04) | 0.61 | 0.62 | <0.01 | 0.89 | <0.01 |
| Majority tribe | 1.08 (0.82, 1.56) | 1.00 | 1.10 | 1.00 | 1.01 | 1.00 |
| Majority religion | 1.00 (0.79, 1.11) | 1.00 | 1.00 | 1.00 | 1.00 | 1.00 |
| Years in education | 1.01 (0.96, 1.08) | 1.00 | 1.02 | 0.21 | 1.01 | 1.00 |
| Farmer | 1.17 (0.78, 1.88) | 1.00 | 0.97 | 1.00 | 0.99 | 1.00 |
| Fisherman | 1.79 (0.91, 4.42) | 0.78 | 2.00 | <0.01 | 1.23 | <0.01 |
| Fishmonger | 1.27 (0.63, 3.73) | 1.00 | 1.58 | 0.12 | 1.09 | 1.00 |
| Home quality score | 1.00 (0.96, 1.06) | 1.00 | 0.99 | 0.23 | 1.00 | 1.00 |
| Household social status | 1.00 (0.76, 1.29) | 1.00 | 1.02 | 1.00 | 1.00 | 1.00 |
| Number of individuals in HH | 1.06 (0.96, 1.22) | 1.00 | 1.09 | <0.01 | 1.02 | 1.00 |
| Years HH lived in village | 1.00 (0.99, 1.00) | 1.00 | 1.00 | 1.00 | 1.00 | 1.00 |
| Home owned | 0.95 (0.50, 1.50) | 1.00 | 1.05 | 1.00 | 1.02 | 1.00 |
| Number of rooms | 0.99 (0.90, 1.05) | 1.00 | 1.00 | 1.00 | 1.00 | 1.00 |
| Current alcohol use | 1.01 (0.80, 1.38) | 1.00 | 1.05 | 1.00 | 1.00 | 1.00 |
| Improved drinking water source | 1.11 (0.78, 1.73) | 1.00 | 0.95 | 0.01 | 1.03 | 0.19 |
| Number of water activities | 1.00 (0.94, 1.07) | 1.00 | 1.00 | 1.00 | 1.00 | 1.00 |
| Year of recruitment - 2023 | 1.00 (0.78, 1.20) | 1.00 | 0.99 | 1.00 | 1.00 | 1.00 |
| Min. dist. (km) to water site | 0.94 (0.61, 1.30) | 1.00 | 0.86 | 0.2 | 1.00 | 0.07 |
| Min. dist. (km) to health centre | 1.01 (0.98, 1.06) | 1.00 | 1.01 | 0.59 | 1.00 | 1.00 |
| Buliisa | 1.03 (0.57, 2.11) | 1.00 | 1.31 | <0.01 | 1.08 | <0.01 |
| Pakwach | 1.43 (0.74, 2.82) | 1.00 | 1.99 | <0.01 | 1.28 | <0.01 |

Table S3: Covariates significance on full population.
GOV: gastro-oesophageal varices, PPF: periportal fibrosis, and overall: all 45 conditions. Significance is determined by $q$-values. For gastro-oesophageal varices, significant covariates are also listed in Table 2, the 95% credible interval is based on the highest posterior density. Significance on periportal fibrosis and overall are computed for grouped outcomes, multiple distributions are combined and potentially multi-modal, therefore the mean are reported instead of medians and credible intervals are not provided as they are not meaningful. Periportal fibrosis combines the densities of the five liver patterns C - F, overall combines all 45 densities. The scales of the odds ratios represented the average per unit rate of each covariate.

| Covariate | GOV Median OR (95% CI) | GOV $q$ | PPF Mean OR | PPF $q$ | Overall Mean OR | Overall $q$ |
|---|---|---|---|---|---|---|
| *S. mansoni* log(epg+1) | 0.93 (0.72, 1.05) | 0.67 | 0.97 | 0.15 | 0.97 | 1.00 |
| Malaria | 0.91 (0.49, 1.34) | 1.00 | 0.86 | 1.00 | 0.86 | 1.00 |
| HBV | 1.31 (0.85, 2.89) | 1.00 | 1.16 | 1.00 | 1.16 | 1.00 |
| HIV | 2.12 (0.98, 6.92) | 0.80 | 1.59 | 0.03 | 1.59 | 1.00 |
| log(Hb concentration) | 0.07 (0.01, 0.37) | <0.01 | 0.66 | 0.01 | 0.66 | <0.01 |
| Age | 1.06 (1.03, 1.10) | 1.00 | 1.05 | <0.01 | 1.05 | <0.01 |
| Gender - female | 0.66 (0.25, 1.04) | 0.09 | 0.61 | <0.01 | 0.61 | <0.01 |
| Majority tribe | 1.08 (0.82, 1.56) | 1.00 | 1.10 | 1.00 | 1.10 | 1.00 |
| Majority religion | 1.00 (0.79, 1.10) | 1.00 | 1.00 | 1.00 | 1.00 | 1.00 |
| Years in education | 1.01 (0.96, 1.07) | 1.00 | 1.02 | 1.00 | 1.02 | 1.00 |
| Farmer | 1.13 (0.82, 1.66) | 1.00 | 0.98 | 0.33 | 0.98 | 1.00 |
| Fisherman | 1.54 (0.93, 3.03) | 1.00 | 1.68 | 0.07 | 1.68 | 1.00 |
| Fishmonger | 1.19 (0.71, 2.60) | 1.00 | 1.39 | 0.88 | 1.39 | 1.00 |
| Home quality score | 1.00 (0.96, 1.06) | 1.00 | 0.99 | 0.23 | 0.99 | 1.00 |
| Household social status | 1.00 (0.76, 1.29) | 1.00 | 1.02 | 1.00 | 1.02 | 1.00 |
| Number of individuals in HH | 1.06 (0.96, 1.22) | 1.00 | 1.09 | 0.03 | 1.09 | 1.00 |
| Years HH lived in village | 1.00 (0.99, 1.00) | 1.00 | 1.00 | 1.00 | 1.00 | 1.00 |
| Home owned | 0.95 (0.51, 1.50) | 1.00 | 1.05 | 1.00 | 1.05 | 1.00 |
| Number of rooms | 0.99 (0.90, 1.05) | 1.00 | 1.00 | 1.00 | 1.00 | 1.00 |
| Current alcohol use | 1.01 (0.84, 1.28) | 1.00 | 1.04 | 1.00 | 1.04 | 1.00 |
| Improved drinking water source | 1.11 (0.78, 1.73) | 1.00 | 0.95 | 0.18 | 0.95 | 1.00 |
| Number of water activities | 1.00 (0.94, 1.07) | 1.00 | 1.00 | 1.00 | 1.00 | 1.00 |
| Year of recruitment - 2023 | 1.00 (0.78, 1.20) | 1.00 | 0.99 | 1.00 | 0.99 | 1.00 |
| Min. dist. (km) to water site | 0.94 (0.61, 1.30) | 1.00 | 0.85 | 0.08 | 0.85 | 1.00 |
| Min. dist. (km) to health centre | 1.01 (0.98, 1.06) | 1.00 | 1.01 | 1.00 | 1.01 | 1.00 |
| Buliisa | 1.03 (0.56, 2.11) | 1.00 | 1.31 | 0.01 | 1.31 | <0.01 |
| Pakwach | 1.43 (0.74, 2.83) | 1.00 | 1.99 | <0.01 | 1.99 | <0.01 |

Table S4: Covariates significance on adults.
GOV: gastro-oesophageal varices, PPF: periportal fibrosis, and overall: all 45 conditions. Significance is determined by $q$-values. For gastro-oesophageal varices, significant covariates are also listed in Table S1, the 95% credible interval is based on the highest posterior density. Significance on periportal fibrosis and overall are computed for grouped outcomes, multiple distributions are combined and potentially multi-modal, therefore the mean are reported instead of medians and credible intervals are not provided as they are not meaningful. Periportal fibrosis combines the densities of the five liver patterns C - F, overall combines all 45 densities. The scales of the odds ratios represented the average per unit rate of each covariate.

| Covariate | GOV Median OR (95% CI) | GOV $q$ | PPF Mean OR | PPF $q$ | Overall Mean OR | Overall $q$ |
|---|---|---|---|---|---|---|
| *S. mansoni* log(epg+1) | 1.04 (0.93, 1.11) | 1.00 | 1.00 | 1.00 | 1.00 | 1.00 |
| Malaria | 1.11 (0.43, 1.80) | 1.00 | 0.97 | 1.00 | 1.02 | <0.01 |
| HBV | 0.72 (0.38, 1.91) | 1.00 | 1.03 | 1.00 | 0.98 | 1.00 |
| HIV | 1.23 (0.57, 2.04) | 1.00 | 1.01 | 1.00 | 1.01 | 1.00 |
| log(Hb concentration) | 0.39 (0.07, 3.76) | 1.00 | 1.40 | 1.00 | 0.57 | <0.01 |
| Age | 1.03 (0.93, 1.20) | 1.00 | 1.04 | 1.00 | 1.02 | <0.01 |
| Gender - female | 0.93 (0.68, 1.57) | 1.00 | 1.04 | 1.00 | 0.96 | 1.00 |
| Majority tribe | 1.00 (0.81, 1.47) | 1.00 | 1.00 | 1.00 | 1.00 | 1.00 |
| Majority religion | 0.88 (0.82, 1.33) | 1.00 | 0.99 | 1.00 | 1.00 | 1.00 |
| Years in education | 0.97 (0.92, 1.07) | 1.00 | 1.00 | 1.00 | 1.00 | 1.00 |
| Farmer | 0.94 (0.37, 3.09) | 1.00 | 1.03 | 1.00 | 0.99 | 1.00 |
| Fisherman | 0.06 (0.01, 2.19) | 1.00 | 0.51 | 1.00 | 0.88 | 1.00 |
| Home quality score | 0.98 (0.97, 1.03) | 1.00 | 1.00 | 1.00 | 1.00 | 1.00 |
| Household social status | 1.22 (0.68, 1.35) | 1.00 | 1.02 | 1.00 | 1.01 | 1.00 |
| Number of individuals in HH | 1.05 (0.95, 1.11) | 1.00 | 1.00 | 1.00 | 1.00 | 1.00 |
| Years HH lived in village | 1.00 (1.00, 1.00) | 1.00 | 1.00 | 1.00 | 1.00 | 1.00 |
| Home owned | 1.06 (0.63, 1.40) | 1.00 | 1.02 | 1.00 | 1.02 | 1.00 |
| Number of rooms | 1.00 (0.94, 1.05) | 1.00 | 1.00 | 1.00 | 1.00 | 1.00 |
| Current alcohol use | 1.24 (0.35, 2.40) | 1.00 | 0.72 | 1.00 | 0.95 | 1.00 |
| Improved drinking water source | 1.23 (0.79, 1.36) | 1.00 | 0.95 | 1.00 | 0.99 | 1.00 |
| Number of water activities | 1.00 (0.94, 1.04) | 1.00 | 1.00 | 1.00 | 1.00 | 1.00 |
| Year of recruitment - 2023 | 1.21 (0.70, 2.12) | 1.00 | 1.01 | 1.00 | 1.01 | 1.00 |
| Min. dist. (km) to water site | 1.36 (0.50, 1.49) | 1.00 | 0.99 | 1.00 | 1.02 | 0.15 |
| Min. dist. (km) to health centre | 1.00 (0.97, 1.05) | 1.00 | 1.02 | 1.00 | 1.00 | 1.00 |
| Buliisa | 1.69 (0.74, 2.35) | 1.00 | 1.02 | 1.00 | 1.01 | 1.00 |
| Pakwach | 1.20 (0.79, 1.44) | 1.00 | 1.07 | 1.00 | 1.03 | 1.00 |

Table S5: Covariates significance on children.
GOV: gastro-oesophageal varices, PPF: periportal fibrosis, and overall: all 45 conditions. Significance is determined by $q$-values. For gastro-oesophageal varices, the 95% credible interval is based on the highest posterior density. Significance on periportal fibrosis and overall are computed for grouped outcomes, multiple distributions are combined and potentially multi-modal, therefore the mean are reported instead of medians and credible intervals are not provided as they are not meaningful. Periportal fibrosis combines the densities of the five liver patterns C - F, overall combines all 45 densities. The scales of the odds ratios represented the average per unit rate of each covariate.

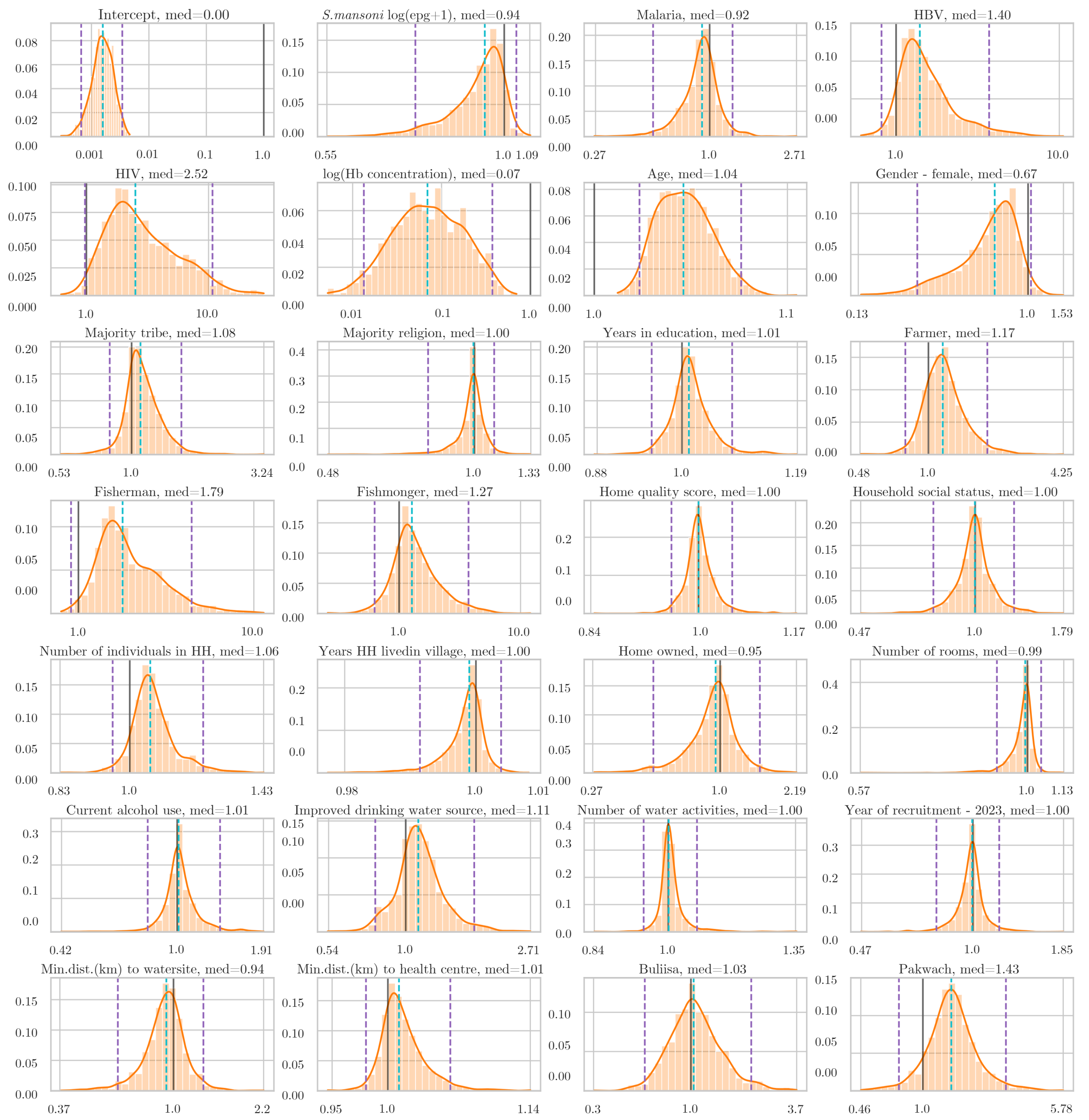


Figure S2: Posterior distribution of multitask regression odds ratios for gastro-oesophageal varices.

The distributions correspond to the column of regression coefficients for gastro-oesophageal varices in Fig. S3. Black lines mark odds ratio 1, cyan dashed lines mark the medians, and purple dashed lines mark the 95% highest posterior density credible intervals.

| Condition | None | Mild | Moderate | Severe | $\chi^2$ | $p_\chi$ |
|---|---|---|---|---|---|---|
| Patches | 48.89 | 13.33 | 26.67 | 11.11 | 6.81 | 0.08 |
| Patches (absent) | 61.90 | 13.79 | 21.32 | 2.99 | | |
| Chronic hepatitis or early cirrhosis | 38.46 | 23.08 | 30.77 | 7.69 | 11.86 | 0.01 |
| Chronic hepatitis or early cirrhosis (absent) | 62.20 | 13.59 | 21.20 | 3.01 | | |
| Left liver lobe moderately enlarged | 55.69 | 16.11 | 22.04 | 6.16 | 2.08 | 0.56 |
| Left liver lobe moderately enlarged (absent) | 62.64 | 13.43 | 21.30 | 2.63 | | |
| Left liver lobe severely enlarged | 51.19 | 14.29 | 25.00 | 9.52 | 4.83 | 0.18 |
| Left liver lobe severely enlarged (absent) | 62.00 | 13.77 | 21.30 | 2.93 | | |
| Caudal liver edge rounded | 45.26 | 18.25 | 28.47 | 8.03 | 7.69 | 0.05 |
| Caudal liver edge rounded (absent) | 63.28 | 13.36 | 20.72 | 2.64 | | |
| Mean portal vein severely enlarged | 53.61 | 15.06 | 21.08 | 10.24 | 5.08 | 0.17 |
| Mean portal vein severely enlarged (absent) | 62.16 | 13.72 | 21.41 | 2.71 | | |
| Gastro-oesophageal varices | 27.27 | 0.00 | 45.45 | 27.27 | 55.38 | $<$0.01 |
| Gastro-oesophageal varices (absent) | 61.83 | 13.84 | 21.31 | 3.02 | | |
| Splenorenal shunt | 15.38 | 23.08 | 38.46 | 23.08 | 50.68 | $<$0.01 |
| Splenorenal shunt (absent) | 61.90 | 13.75 | 21.32 | 3.02 | | |
| Ascites | 25.00 | 0.00 | 33.33 | 41.67 | 65.68 | $<$0.01 |
| Ascites (absent) | 61.85 | 13.84 | 21.35 | 2.96 | | |
| Spleen length severely enlarged | 35.56 | 18.89 | 36.11 | 9.44 | 16.7 | $<$0.01 |
| Spleen length severely enlarged (absent) | 63.29 | 13.48 | 20.50 | 2.72 | | |
| Spleen length moderately enlarged | 49.74 | 16.41 | 28.12 | 5.73 | 4.2 | 0.24 |
| Spleen length moderately enlarged (absent) | 63.37 | 13.42 | 20.46 | 2.74 | | |

Table S6: Anaemia categories for conditions significant with Hb concentration. Conditions were taken from Table 2. Anaemia category percentages were calculated for each condition over the positive and negative participants separately. Conditions followed by (absent) refer to participants that did not have the condition. $\chi^2$ test is calculated between the percentages of the positive and negative participants of that condition.

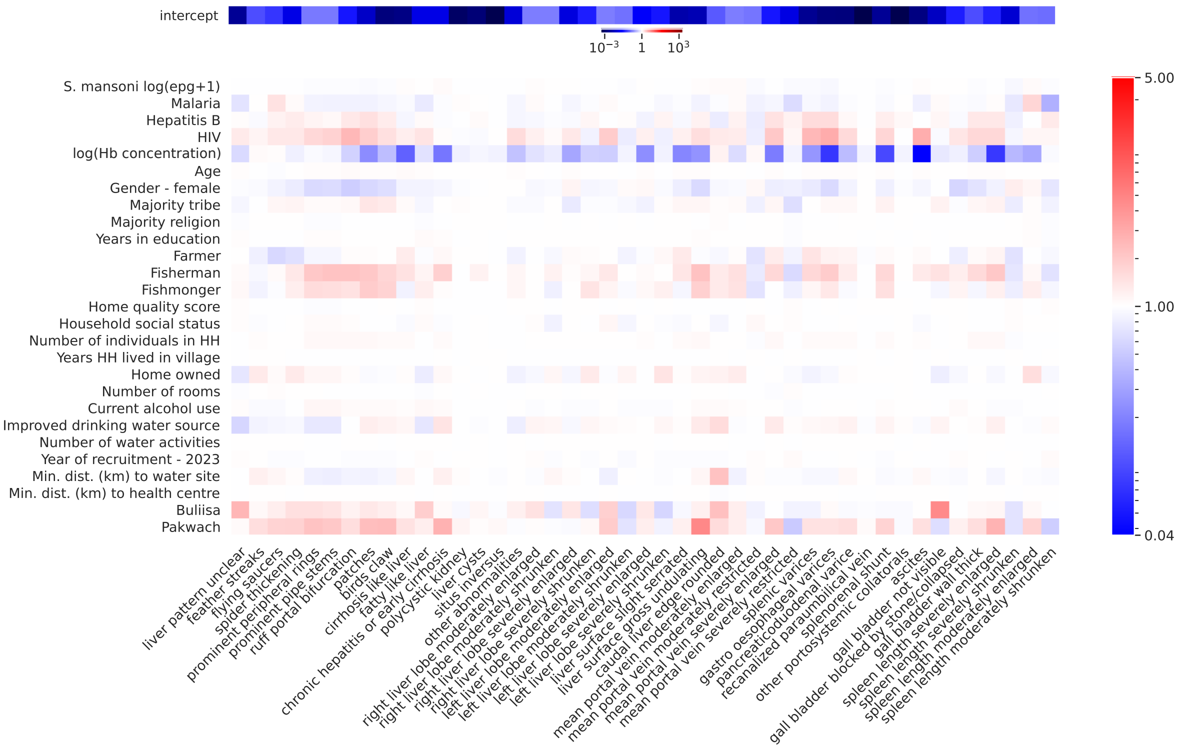


Figure S3: Mean posterior odds ratios of $m_{ij}/\sigma_j$.
Each column shows to the mean odds ratios of the covariates for predicting the corresponding condition of that column.

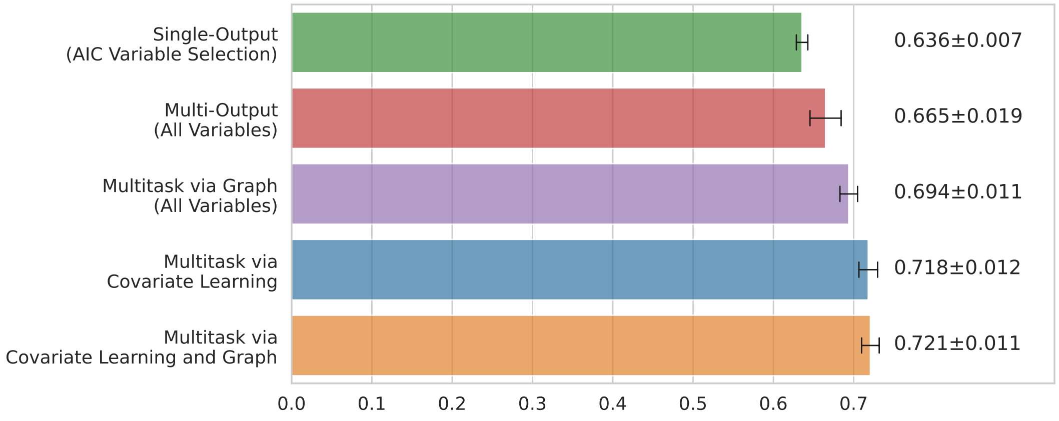


Figure S4: Comparison between different models and frameworks.
The figure shows results comparing multitask models against different ways of modelling multiple outcomes. Green: 45 logistic regression models, each model used individual variable selection based on AIC. Red: Multi-output model and using all variables (no variable selection). Purple: Multi-output model via graph using all variables (no variable selection). Blue: Multitask model via variable selection. Orange: Full multitask model. For green and orange models, AUC for each condition can be found in Fig. 2.

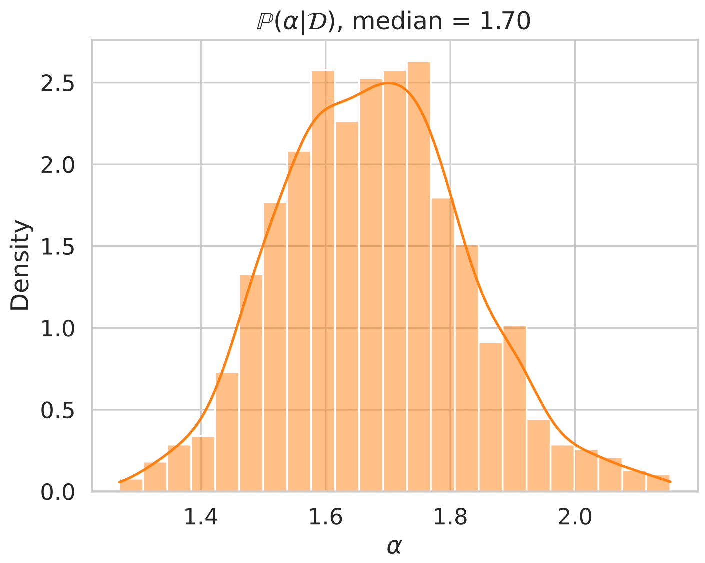


Figure S5: Posterior distribution of $\alpha$ from multitask model.

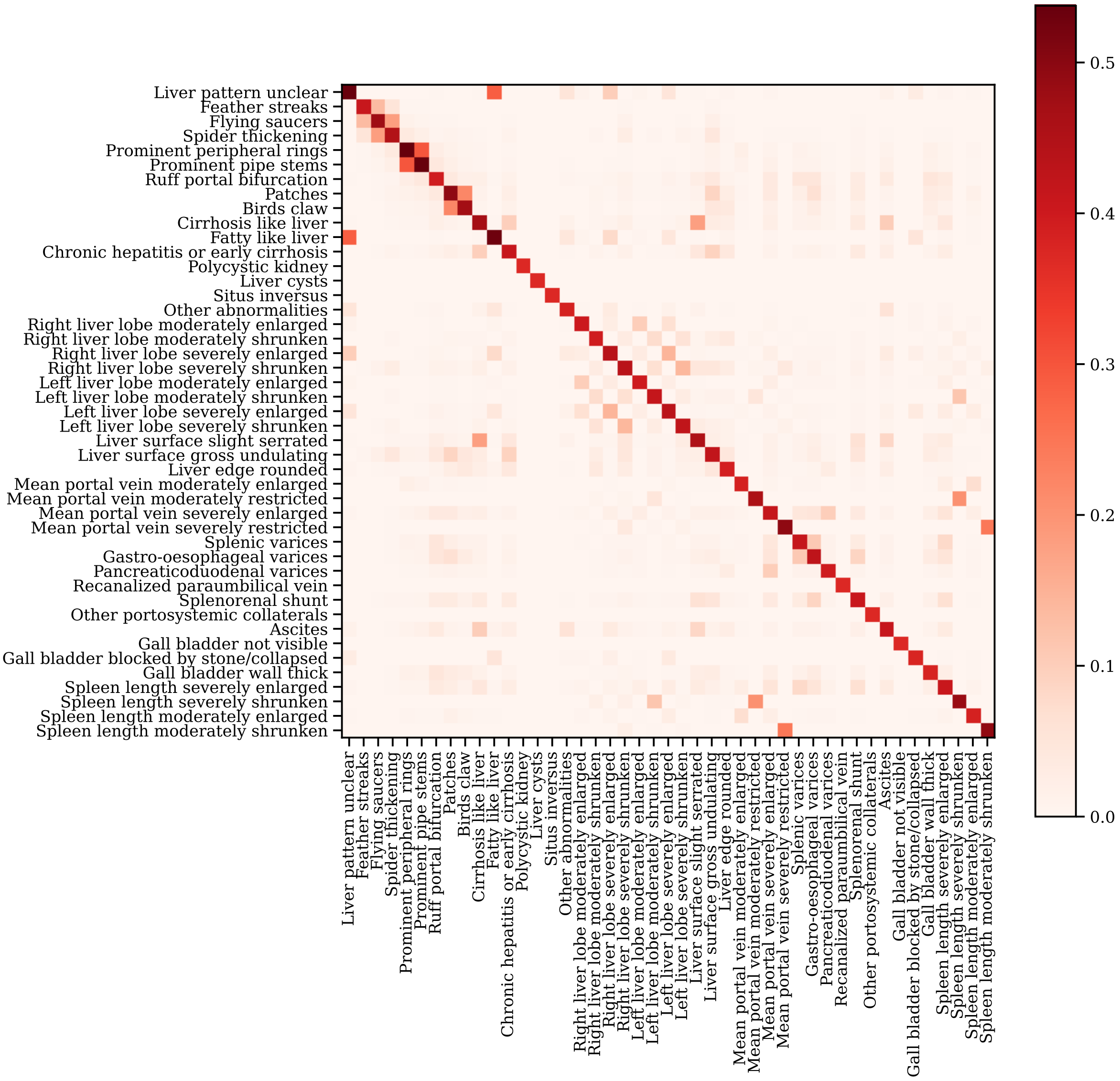


Figure S6: Relative probabilities between all conditions.
Probabilities are determined by the graph convolution matrix $f_\alpha(\mathcal{G})$ using median $\alpha$ - see Fig. S5. Diagonal indicates self influence.

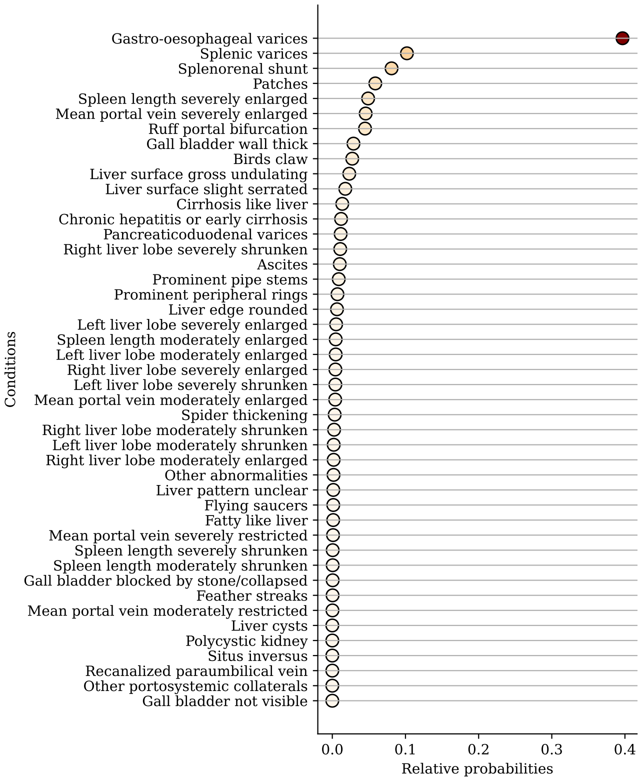


Figure S7: Influence probabilities for gastro-oesophageal varices.

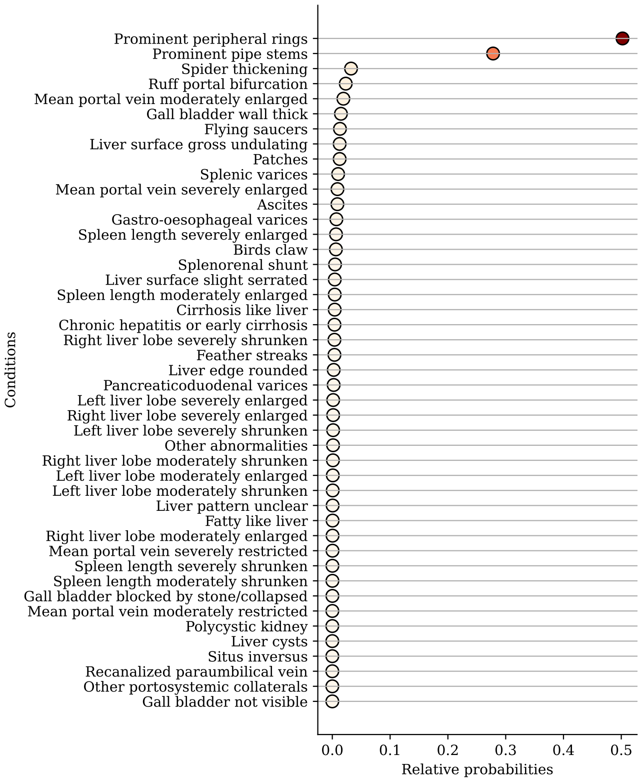


Figure S8: Influence probabilities for prominent peripheral rings liver pattern, grade C1.

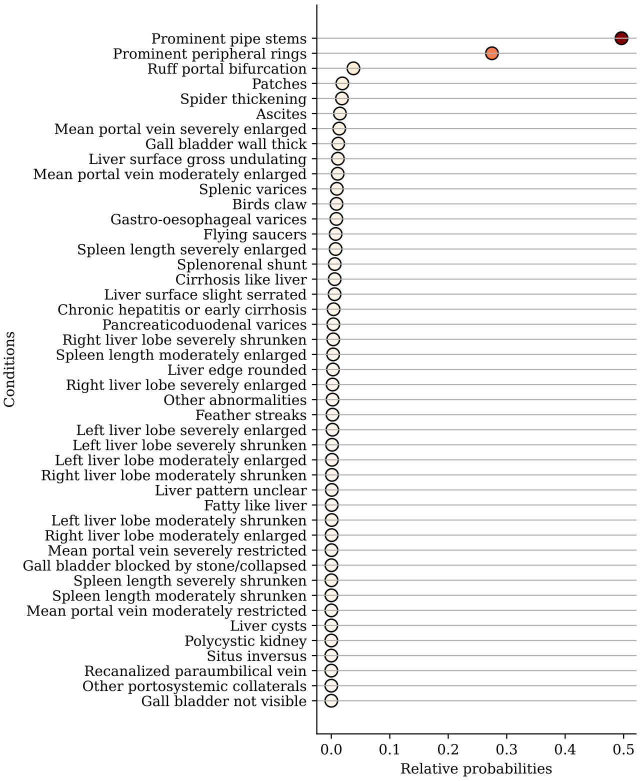


Figure S9: Influence probabilities for prominent pipe stems liver pattern, grade C2.

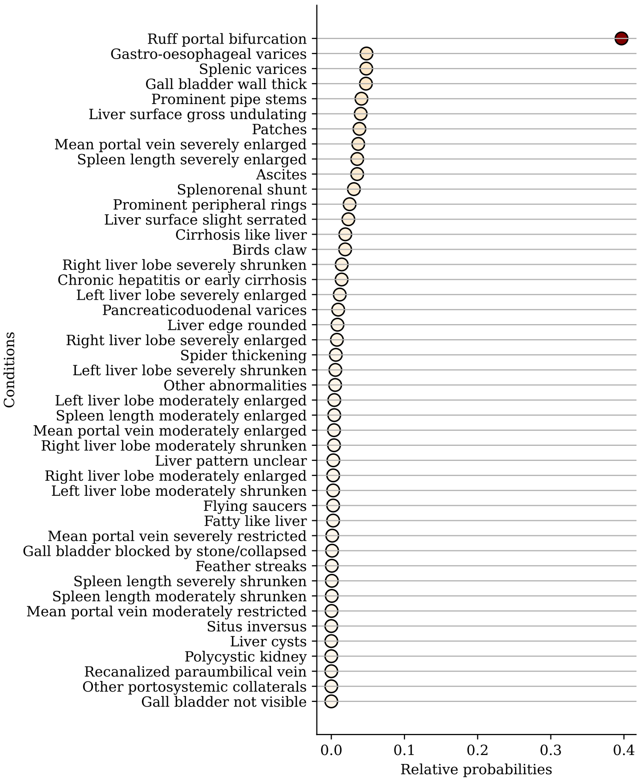


Figure S10: Influence probabilities for ruff portal bifurcation liver pattern, grade D.

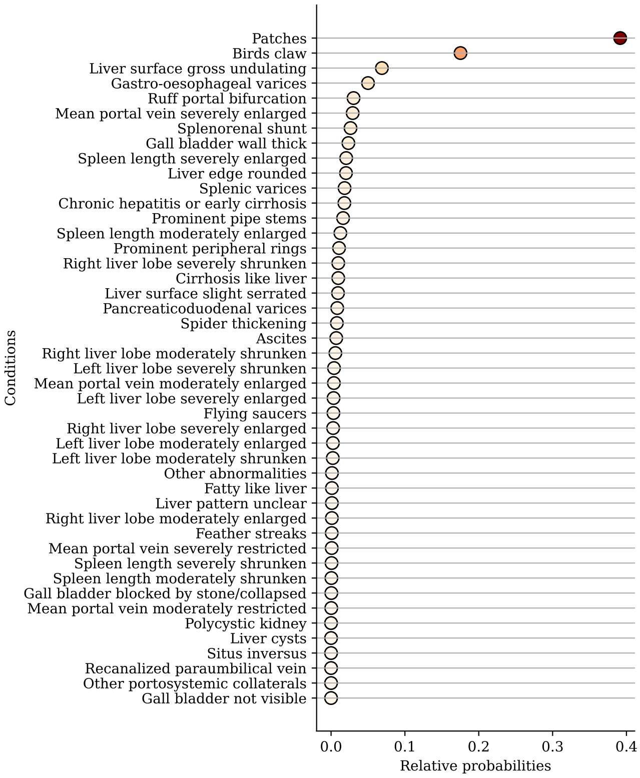


Figure S11: Influence probabilities for patches liver pattern, grade E.

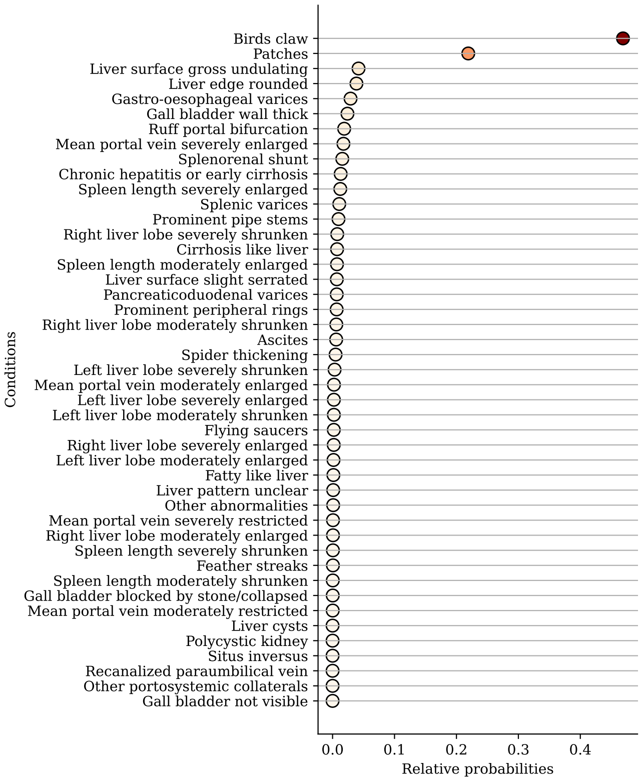


Figure S12: Influence probabilities for birds claw liver pattern, grade F.
